# A meta-analysis of bone conduction 80 Hz auditory steady state response thresholds in adults and infants

**DOI:** 10.64898/2026.02.12.26346168

**Authors:** Emanuele Perugia, Constantina Georga

## Abstract

**Background:** Auditory steady-state responses (ASSRs) provide an objective method for estimating hearing thresholds in individuals unable to provide behavioural responses. Bone conduction (BC) testing is required to differentiate conductive from sensorineural hearing loss. Accurate BC ASSR threshold estimation relies on “correction” factors, which are not yet well established. This meta-analysis evaluated the reliability of BC ASSR thresholds to estimate hearing thresholds at 500, 1000, 2000 and 4000 Hz.

**Methods:** A systematic search of PubMed, the Cochrane Library, and Embase was conducted to identify studies involving normal-hearing (NH) and hearing-impaired (HI) participants of all ages. Outcomes were (1) the difference between ASSR behavioural and ASSR thresholds, and (2) ASSR thresholds. The risk of bias was evaluated using the Newcastle-Ottawa Scale. The mean and 95% confidence intervals (CI) were calculated for the thresholds at the four frequencies. The certainty of the evidence was assessed using GRADE approach.

**Results:** Of records identified, 12 records met the inclusion criteria, yielding a total of 27 studies. Sample sizes ranged from 60 to 271 participants across frequencies and age groups. The quality of records ranged from low to high. Data were synthesised using random-effects models due to heterogeneity. In NH adults, the mean differences (±95% CI) between BC ASSR thresholds and behavioural thresholds were 17.0 (±4.8), 15.5 (±6.0), 13.4 (±3.3), and 12.1 (±4.1) dB at 500, 1000, 2000, and 4000 Hz, respectively. In NH infants, mean (±95% CI) BC ASSR thresholds were 17.2 (±2.0), 10.5 (±3.6), 26.1 (±2.6), and 19.9 (±4.0) dB HL at the same frequencies. In infants with conductive HL, BC ASSR threshold was 20.3 (±4.6) at 500 Hz. The certainty of the evidence was very low.

**Conclusions:** BC ASSR can be a reliable method for estimating BC thresholds. However, age and frequency significantly impact BC ASSR thresholds, highlighting the need to develop of “correction” factors to accurately predict BC behavioural thresholds.

**Registration:** PROSPERO CRD42023422150.

## Introduction

Auditory Steady-State Responses (ASSRs) are auditory evoked potentials that are used in clinical audiology to estimate hearing thresholds [1–6]. Clinically, the technique is applicable for populations that are not able or not willing to provide behavioural thresholds, such as infants, children, and adults with intellectual difficulties or those with non-organic functional hearing loss.

The main advantage of the ASSR is that it allows objective simultaneous threshold measurements at several frequencies, reducing the testing time required [7]. A second advantage is that interpretation is primarily objective, as it relies on statistical analysis of the signal in the frequency domain [8–10], whereas interpretation of Auditory Brainstem Response (ABR) is dependent primarily on subjective interpretation of the waveform in the time domain [6,7]. The ASSR method is limited, in that, as the response is analysed in the frequency domain, it does not provide information regarding neural integrity, and so cannot diagnose neuropathy [11]. For this reason, the UK clinical ASSR protocol recommends ABR testing as the starting point for neonatal assessments [12]. For a comprehensive discussion of the strengths and weaknesses of ASSR, please refer to [6,13,14].

Bone conduction (BC) testing is important to differentiate between conductive and sensorineural hearing losses which sequentially informs the decisions around the urgency and type of intervention. However, some information about conductive hearing loss can also be obtained through the click-ABR [15], as well as tympanometry and acoustic immittance [16,17]. In mixed hearing losses, BC levels inform the gain set on the hearing aids and thus ensure there is no under- or over-amplification [18]. This is particularly important for infants, where early intervention is crucial and obtaining outcome measures on hearing aid fitting is most difficult. Although there has been a substantial body of research on air conduction (AC) ASSRs [14], a relatively smaller number of studies have focused on BC ASSR testing, especially when considering normative data for threshold estimation. In general, considerable heterogeneity exists in methods among ASSR studies [14,19] and no universal testing protocol has been established [12,20,21].

The objective of the meta-analysis was to assesses whether hearing thresholds estimated by BC ASSR are reliable at the four classic audiometric frequencies (i.e., 500, 1000, 2000 and 4000 Hz). The present study sought to synthesise the BC ASSR thresholds from studies conducted to date. Research in clinical auditory electrophysiology generally consists of observational studies Our starting intention was to collect data from all ages. Data from infants are very limited with many studies demonstrating methodological issues, such as different procedures to record BC ASSR [22,23], lack of behavioural BC thresholds for comparison and frequency-specific hearing screening [24].

## Methods

The study was pre-registered in PROSPERO (CRD42023422150) and followed the PRISMA 2020 statement [25,26].

### Literature search, data collection, and risk of bias

#### Eligibility criteria

Eligibility criteria for study characteristics were based on the Population, Interventions, Comparators and Outcomes (PICO) framework [27]. Population: both normal-hearing (NH) and hearing-impaired (HI) participants of all ages. Interventions: BC ASSR thresholds. Comparators: BC ASSR thresholds vs. BC behavioural thresholds in adults and infants, and in NH and HI participants. Outcomes: the main outcomes were the BC ASSR thresholds in NH participants, as well as the difference between BC behavioural and ASSR thresholds. Additional (co-)factors were participants’ age and hearing threshold, testing levels and frequencies.

Only studies that reported BC ASSR thresholds were included. Studies were excluded if they met any of the following criteria: (1) not published in English, (2) pre-prints (i.e., not peer-reviewed), (3) reviews, (4) animal studies, (5) BC ASSR data collected from participants with a genetic syndrome or condition, (6) BC ASSR thresholds not reported, (7) BC ASSRs recorded in cochlear implant or active middle ear implant users, (8) BC ASSRs evoked by a 40 Hz modulation rate, or (9) unavailable as a full-text version. BC ASSRs evoked by a 40 Hz modulation rate were excluded because ASSRs evoked by an 80 Hz modulation rate are recommended for assessing hearing thresholds [12], and have different neural generators and amplitude than those evoked by a 40 Hz modulation rate [13,28,29]. No restrictions were applied regarding the date of publication.

#### Information sources and search strategy

The MEDLINE, Embase, and Cochrane Central Register of Controlled Trials (CENTRAL) databases were searched by the authors from May 2023 to August 2023 (see Supplementary Table S1 for the full search strategies). Using the same strategy, a further search was conducted in December 2025 to update (and verify) the initial search. Additionally, backward and forward citation searchings were performed using the *Citationchaser* Shiny app [30]. Specifically, all references of the included reports (backward search) and all studies that cited the most recent included report (forward search) were scanned.

#### Selection process

The authors independently screened titles and abstracts of potentially eligible studies. Then, they independently reviewed full-text articles for inclusion. In both stages, disagreements between authors were solved by discussion.

#### Data collection process

EP extrapolated data items and entered them into a spreadsheet. CG checked data accuracy. Disagreement was solved by re-checking articles. Data collection was performed by entering values from tables in the source articles, by synthesis of values from figures in the source articles via WebPlotDigitizer [31] (i.e., Fig 3 of [24] and Fig 3 of [32]), or by a table from the related dissertation (i.e., data of [33] were taken from [34]).

#### Data items

Data and information collected from the studies were the following: study (authors, year, and country), participants (sample size, age, and hearing threshold and group), equipment (device, bone conductor type and position, electrode montage), protocol (stimuli and their calibration, artefact rejection, average type, detection algorithm, noise criterion, and testing frequencies and levels). Missing data are identified and reported.

#### Study risk of bias assessment

The Newcastle-Ottawa Scale (NOS) for cohort studies [35] was used to assess the risk of bias in the included studies. The assessment covered three categories (Selection, Comparability, and Outcome) and collectively identified over eight items. Items in the Selection and Outcome categories could be awarded a maximum of one star each, while the item in the Comparability category could receive up to two stars. Studies with 0-3 stars were considered high risk, those with 4-6 stars intermediate risk, and those with 7-9 stars low risk [36]. As stated in the pre-registration, a minimum of three papers with at least three stars on the NOS tool was required for inclusion in the synthesis. No sensitivity analyses were planned. The two authors independently assessed the studies and resolved any discrepancies through discussion.

### Effect measures, synthesis methods, and certainty assessment

#### Effect measures

Analyses were performed on the BC ASSR thresholds for NH participants, and on the difference between their behavioural and ASSR BC thresholds. The thresholds were analysed as a function of participants’ age and testing frequency. Mean, SD, effect size and 95% confidence intervals (CIs) were calculated. Due to the insufficiency of reported results for HI participants, analyses of the BC ASSR thresholds for these participants could not be performed.

#### Synthesis methods

The scope of the synthesis was twofold. First, it aimed to compare BC behavioural and ASSR thresholds for both NH and HI participants, with the goal of establishing correction factors for accurately predicting BC behavioural thresholds from BC ASSR thresholds. Second, it involved pooling BC ASSR thresholds for NH participants to establish typical BC ASSR thresholds with the aim of using these as clinical discharge criteria.

However, only three studies (two in adults and one in infants) reported results for HI participants. In general, most articles reported only ASSR BC thresholds, i.e., behavioural thresholds were missing. Given the thresholds variability associated with age and frequency, data subgroups were made based on participants’ age (i.e., adults or infants) and on frequency (i.e., 500, 1000, 2000, and 4000 Hz).

To assess statistical heterogeneity of the data, the I^2^ index [37] was performed within each subgroup. Specifically, the I^2^ index measures the proportion of total variability due to between-study heterogeneity [38]. Either fixed-effect modelling (in case of I^2^ < 50%; i.e., low heterogeneity) or random-effect modelling (otherwise) are used to synthesise the data. The I^2^ index is based on the Chi-squared test, with a statistically significant (i.e., between-study heterogeneity) *p*-value threshold of < 0.10 [37]. In the fixed-effect model, the heterogeneity is neglected and the difference among studies is assumed as being due to chance. Instead, in the random-effect model, the heterogeneity is incorporated.

The objective of the meta-analysis was to assess whether hearing thresholds estimated by BC ASSR are reliable at the four classic audiometric frequencies (i.e., 500, 1000, 2000 and 4000 Hz) and for both adults and infants. To be considered a reliable threshold, the 95% confidence intervals around the mean of the synthesised data should be within 10 dB, as BC ASSR stimuli are generally presented in 10 dB steps [22].

All analyses were run in R (version 4.3.1; [39]) using the packages *PRISMA2020* [40], *meta* [41], and the *Tidyverse* family [42].

#### Certainty assessment

The GRADE approach was used to assess the certainty of the evidence. As the reports included in the meta-analysis were non-randomised studies, by definition, the body of evidence initially began with a low-certainty rating. The following criteria were considered for downgrading the certainty of evidence, where appropriate: unexplained heterogeneity or inconsistency of results, indirectness of evidence, imprecision of results, and a high probability of publication bias. The certainty of evidence could be upgraded in the presence of a large effect [43]. The GRADEpro GDT software was used to prepare the Summary of Findings tables.

## Results

### Study selection

The database searches identified 98 records. After removal of duplicates, 59 records were screened. Twenty-seven records were excluded because they were not in English, were preprints or conference abstracts, were reviews, were not relevant, involved genetic conditions, or involved participants with cochlear implants or active middle ear implants. Three records were not available. Consequently, 29 records underwent full-text review. Of these, 20 records were excluded because they did not performed BC ASSR thresholds, did not report BC ASSR thresholds, reported dB values that could not be converted, or were conducted at 40 Hz. Citation searching of the included records identified 265 additional records, which were scanned, and two met the final selection criteria. After this work was available as a preprint in medRxiv, Professor Emeritus David R. Stapells emailed one of the authors to inform them of a further record, Valeriote & Small 2024 [44], which had been identified through database searches and was excluded as preprint, but has since been published. Therefore, in total, 12 records were remained, which included 29 individual studies [22–24,32,33,44–50]. A summary of the selection procedure is presented in Fig. 1.

**Figure 1:**
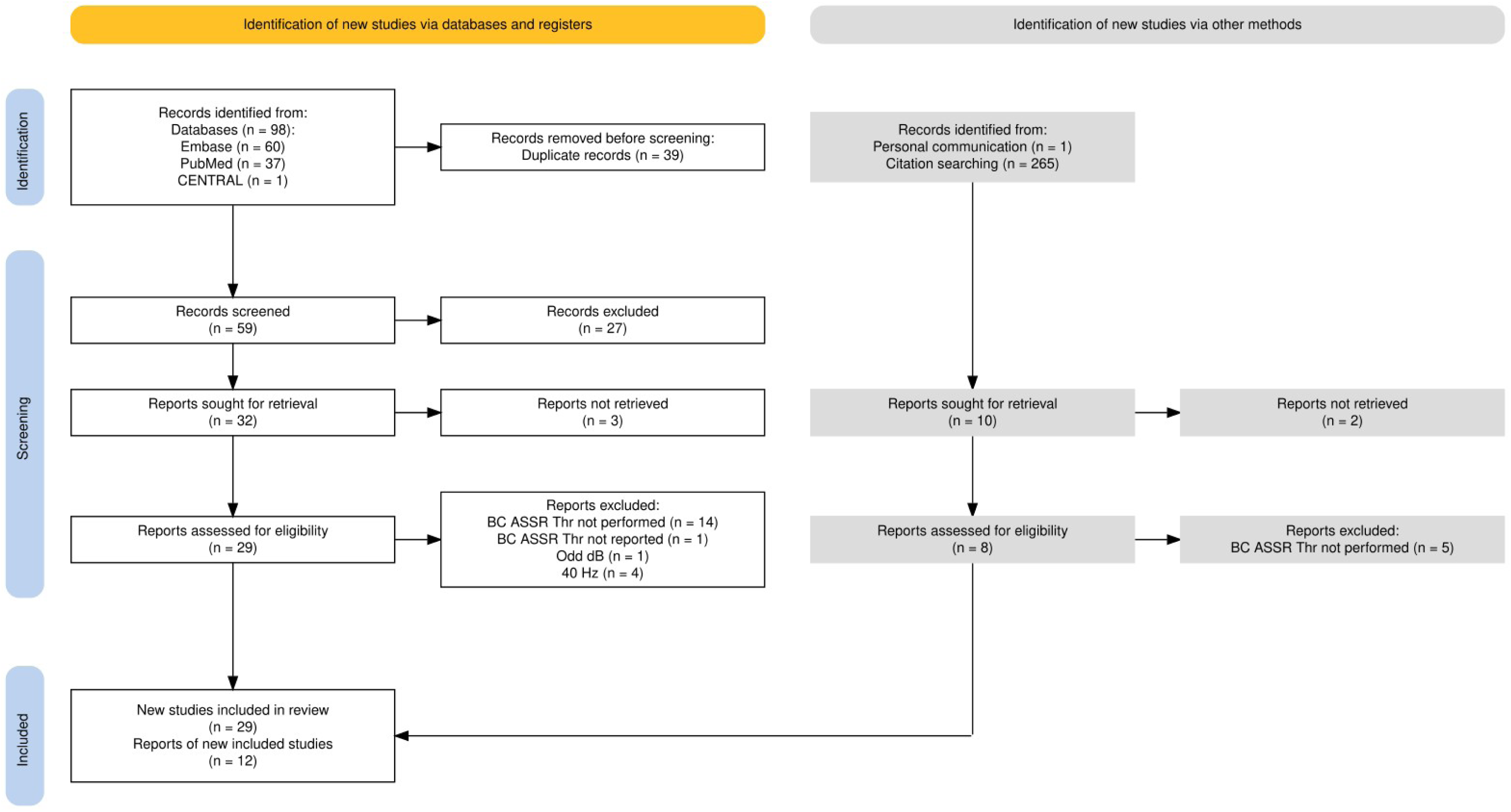
PRISMA 2020 flow diagram of the study selection procedure.

The number of studies was larger than the number of reports, as several reports tested different groups of participants. Specifically, Small & Stapells (2006) [46] measured thresholds in both preterm and post-term infants. Small et al. (2007) [23] reported three different experiments involving three different groups of infants. Small & Stapells (2008a) [24] had three different age groups. Small & Stapells (2008b) [47] and Casey & Small [33] recorded thresholds in infants and adults. Swanepoel et al. (2008) [48] measured thresholds in NH infants and in three groups of HI infants (conductive HL, mild-to-moderate and severe-to-profound sensorineural hearing loss [SNHL]). Ishida et al (2011) [49] tested two groups of NH adults and a group of HI adults. Small & Hu (2011) [32] reported thresholds obtained in young infants, older infants, and adults. Ismaila et al., [50] measured thresholds in NH infants and in two groups of HI infants (mild-to-moderate and severe-to-profound SNHL). Valeriote & Small measured thresholds in NH infants with NH and conductive HL [44].

### Study characteristics

The characteristics of the 12 included reports are presented in Tables 1 and 2. The Correspondent author on eight out of 11 reports were based at the University of British Columbia (Vancouver, Canada) as the group led by David R. Stapells and Susan A. Small have produced many contributions in the field [22–24,32,33,46,47]. The other reports were based at the University of Toronto (Canada) [45], University of Pretoria (South Africa) [48], Al-Azhr University (Cairo, Egypt) [50], and Royal University Hospital (Saskatoon, Canada) [44] with Susan A. Small as co-author. All reports tested participants within different age groups (e.g., adults and infants) and/or different hearing groups (e.g., NH and HI).

**Table 1:** Study Characteristics of the 11 Included Studies. AABR: automatic auditory brain stem response; AC: air-conduction thresholds; BC: bone-conduction thresholds; DPOAE: distortion-product otoacoustic emissions; HI: hearing-impaired participants; NA: not available; NH: normal-hearing participants; PCA: post-conceptional age; SNHL: sensorineural hearing loss; TEOAE: transient evoked otoacoustic emission.

| Study | Country | Sample | Age (Mean) | Hearing status | Hearing Estimation |
| --- | --- | --- | --- | --- | --- |
| Lins et al (1996) [45] | Canada | 8 adults | NA | NH | Behavioural AC & BC @ 500 to 4000 Hz |
| Small & Stapells (2005) [22] | Canada | 10 adults | Range : 18-48 years | NH | Behavioural AC & BC ≤ 20 dB HL @ 250 to 8000 Hz |
| Small & Stapells (2006) [46] | Canada | 29 preterm infants | 34.5 week PCA | NH | AABR at 35 dB nHL |
|  |  | 14 postterm infants | 17 week | NH | DPOAE |
| Small et al (2007) [23] | Canada | 10 postterm infants | 17 week | NH | AABR at 35 dB nHL or DPOAE |
|  |  | 15 preterm infants (9 were included in the analysis of ASSR threshold) | 34.5 week PCA | NH | AABR at 35 dB nHL |
|  |  | 13 postterm infants | 15 week | NH | DPOAE |
| Small & Stapells (2008a) [24] | Canada | 35 young infants (but 9 from [23,47]; 14 from [46]; 12 from <sup>a</sup>) | 16 week | NH |  |
|  |  | 13 older infants | 18.2 months | NH | AABR at 35 dB nHL or DPOAE |
| | | 18 adults (but 10 from [22]) | 22.9 years | NH | Behavioural AC & BC thresholds ≤ 25 dB HL @ 500-4000 Hz |
| Small & Stapells (2008b) [47] | Canada | 14 infants (with 1 participated also in [23]) | 21 week | NH | DPOAE |
| | | 11 adults | 23 years | NH | Behavioural AC thresholds ≤ 25 dB HL @ 500-4000 Hz |
| Swanepoel et al (2008) [48] | South Africa | 13 infants (21 ears) | 3.6 years | NH | Tympanograms, DPOAE, AC click-evoked ABR thresholds, AC ASSR thresholds |
|  |  | 18 infants (35 ears) | 2.7 years | Conductive HL |  |
|  |  | 7 infants (13 ears) | 2.5 years | Mild-to-Moderate SNHL |  |

<sup>a</sup> The manuscript referred to “Small & Stapells (2005b)”, but this was not in the reference of the report. We think that the correct reference is Small & Stapells (2008b) because the number of infants tested using a two-channel EEG.

|  |  |  |  |  |  |
| --- | --- | --- | --- | --- | --- |
|  |  | 13 infants (23 ears) | 2.2 years | Severe-to-profound SNHL |  |
| Ishida et al (2011) [49] | Canada | 16 adults | 26.8 years | NH | Behavioural AC thresholds $\leq 20$ dB HL @ 500-4000 Hz |
|  |  | 10 adults | 24.6 years | NH |  |
| | | 40 adults | 46.0 years | SNHL | Behavioural AC thresholds $> 20$ & $\leq 55$ dB HL @ 500-4000 Hz; & BC thresholds |
| Small & Hu (2011) [32] | Canada | 22 young infants (but for 12 from [23]) | 2.1 months | NH | TEOAE |
|  |  | 10 older infants | 15.3 months | NH |  |
| | | 20 adults | 26.5 years | NH | Behavioural AC & BC thresholds $\leq 25$ dB HL @ 500-4000 Hz |
| Casey & Small (2014) [33] | Canada | 23 infants | 10.5 months | NH | TEOAE or tympanometry |
| | | 12 adults | 29.9 years | NH | Behavioural AC & BC thresholds $\leq 25$ dB HL @ 500-4000 Hz & air-bone gaps $\leq 10$ dB |
| Ismaila et al (2016) [50] | Egypt | 20 infants (35 ears) | 4.5 years | NH | AC play audiometry mean = 15 dB HL, AC click-evoked ABR thresholds mean = 30 dBnHL, tympanograms |
|  |  | 20 infants (36 ears) |  | Mild-to-moderate SNHL | AC & BC play audiometry mean = 30-50 dB HL, AC click-evoked ABR thresholds mean = 40-60 dBnHL, tympanograms |
|  |  | 20 infants (35 ears) |  | Conductive HL | AC play audiometry mean = 25-50 dB HL, Normal BC play audiometry, AC click-evoked ABR elevated, tympanograms |
| Valeriotte & Small (2024) [44] | Canada | Up to 23 ears | 7.36 weeks | NH | AC & BC ABR, TEOAE & tympanograms |
|  |  | 15 ear | 6.71 weeks | Conductive HL | AC & BC ABR, TEOAE & tympanograms |

**Table 2:** Hearing Test Information of the 11 Included Studies AM: amplitude modulation; CR: clear response; CF: carrier frequency; FM: frequency modulation; MF: modulation frequency; HL: hearing loss; HT2: Hotelling T2; PC2: phase coherence squared; RA: response absent; SNHL: sensorineural hearing loss.

| **Study** | **Equipment** | **Placement** | **Stimulus** | **Sweep** | **Electrode montage** | **Artefact rejection** | **Average** | **Detection algorithm (CR)** | **Noise criterion (RA)** | **Testing levels** |
| --- | --- | --- | --- | --- | --- | --- | --- | --- | --- | --- |
| Lins et al (1996) [45] | NA | Forehead | Modulated tones with 100% AM Multiple CF: 0.5, 1, 2, & 4 kHz MF: 77, 85, 93, & 101 Hz | 16 epochs of 512 data points and lasted a total of 12.06 seconds. | Cz and at the nape of the neck | Epoch > $\pm 40$ uV in amplitude rejected. | Normal | F ratio & HT<sup>2</sup> with $p < 0.05$ | 64 sweeps were not SIG | Started at 50 dB SPL, decreased in 10 dB step until all responses were absent |
| Small & Stapells (2005) [22] | Rotman MASTER research system | Temporal Bone | Modulated tones with 100% AM Multiple CF: 0.5, 1, 2, & 4 kHz MF: 77, 85, 93, & 101 Hz | 16 epochs of 1024 data points and lasted a total of 13.107 seconds. | Noninverting on Cz. Inverting at the nape of the neck, just below the hairline. Ground on high forehead. | Epoch > $\pm 40$ uV in amplitude rejected. | Normal | F ratio with $p < 0.05$ | noise < 11 nV & $p > 0.05$ OR noise < 10 nV & $p \geq 0.30$ | 10 dB steps at: (1) 50 to 10 dB HL for noninverted stimuli, (2) 50 to 30 dB HL for inverted stimuli, and (3) 50 to 20 dB HL for alternated stimuli. |
| Small & Stapells (2006) [46] | Rotman MASTER research system | Temporal Bone | Modulated tones with 100% AM & 25% FM Multiple CF: 0.5, 1, 2, & 4 kHz MF: 77, 85, 93, & 101 Hz | 16 epochs of 1024 data points and lasted a total of 13.107 seconds. A minimum of 7 sweeps per condition. | Noninverting on midline at the high forehead. Inverting at the nape of the neck, just below the hairline. Ground at the low forehead. | Epoch > $\pm 40$ uV in amplitude rejected. | Normal | F ratio with $p < 0.05$ for at least two consecutive sweeps. | noise < 11 nV & $p > 0.05$ OR noise < 10 nV & $p \geq 0.30$ | |
| Small et al (2007) [23] | Rotman MASTER research system | Temporal Bone | Modulated tones with 100% AM & 25% FM Multiple CF: 0.5, 1, 2, & 4 kHz MF: 77, 85, 93, & 101 Hz | 16 epochs of 1024 data points and lasted a total of 13.107 seconds. A minimum of 7 sweeps per condition. | Noninverting on forehead. Inverting at the nape of the neck, just below the hairline. Ground on high forehead. | Epoch > $\pm 40$ uV in amplitude rejected. | Weighted | F ratio with $p < 0.05$ for at least two consecutive sweeps. | noise < 11 nV & $p > 0.05$ OR noise < 10 nV & $p \geq 0.30$ | 10-dB steps from 50 to -10 dB HL |
| Small & Stapells (2008a) [24] | Rotman MASTER research system | Temporal Bone | Modulated tones with 100% AM & 25% FM Multiple CF: 0.5, 1, 2, & 4 kHz MF: 77, 85, 93, & 101 Hz | 16 epochs of 1024 data points and lasted a total of 13.107 seconds. A minimum of 7 sweeps per condition. | For 23 of the 35 young infants, all older infants and adults: Noninverting on forehead. Inverting at the nape of the neck. Ground on high forehead. For 12 of the 35 young infants, 2 2 inverting on the L & R mastoids | Epoch > $\pm 40$ uV in amplitude rejected. | Weighted | F ratio with $p < 0.05$ for at least two consecutive sweeps. | noise < 11 nV & $p > 0.05$ OR noise < 10 nV & $p \geq 0.30$ | 10-dB steps |
| Small & Stapells (2008b) [47] | Rotman MASTER research system | Temporal Bone | Modulated tones with 100% AM & 25% FM Multiple CF: | 16 epochs of 1024 data points and lasted a total of 13.107 | Noninverting on high forehead. Inverting on mastoids. | Epoch > $\pm 40$ uV in amplitude rejected. | Weighted | F ratio with $p < 0.05$ for at least two consecutive sweeps. | noise < 11 nV & $p > 0.05$ OR noise < 10 nV & $p \geq$ | 10-dB steps from 50 to -10 dB HL |
|  |  |  | 0.5, 1, 2, & 4 kHz MF: 77, 85, 93, & 101 Hz | seconds. A minimum of 7 sweeps per condition. | Ground on low forehead. |  |  |  | 0.30 |  |
| Swanepoel et al (2008) [48] | Gsi AUDERA | Mastoid | Modulated tones with 100% AM & 10% FM Multiple CF: 0.25, 0.5, 1, 2, & 4 kHz MF: 67, 74, 81, 88, & 95 Hz | NA | Noninverting on high forehead (Fz). Inverting on ipsilateral mastoid. Ground on contralateral mastoid. | | | PC<sup>2</sup> with $p < 0.03$ | 1uV | 10- & 5- dB steps |
| Ishida et al (2011)[49] | Rotman MultiMASTER | Temporal Bone | Modulated tones with 100% AM & 25% FM Multiple CF: 0.5, 1, 2, & 4 kHz MF: 77, 85, 93, & 101 Hz | 16 epochs of 1024 data points and lasted a total of 13.107 seconds. A min of 12 sweeps per condition. | Noninverting on Cz. Inverting on midline at the nape. Ground on high forehead. | Epoch > $\pm 57$ uV in amplitude rejected. | Weighted | F ratio with $p < 0.05$ for at least 7 consecutive sweeps. | noise1 < 11 nV & $p > 0.05$ OR ASSR amp < 10 nV & $p \geq 0.30$ | Started at 20 or 40 dB HL, increased or decreased in 10-dB steps |
| Small & Hu (2011) [32] | Rotman MultiMASTER | Temporal Bone | Modulated tones with 100% AM & 25% FM Multiple CF: 0.5, 1, 2, & 4 kHz MF: 77, 85, 93, & 101 Hz | 16 epochs of 1024 data points and lasted a total of 13.107 seconds. A min of 10 sweeps per condition. | Noninverting on high forehead. Inverting on ipsilateral mastoids. Ground on forehead. | Epoch > $\pm 80$ uV in amplitude rejected. | Weighted | F ratio with $p < 0.05$ for at least 2 consecutive sweeps. | noise < 11 nV & $p > 0.05$ OR ASSR amp < 10 nV & $p \geq 0.30$ | Random start between 10 to 30 dB HL, increased or decreased in 10-dB steps |
| Casey & Small (2014) [33] | Rotman MultiMASTER | Temporal Bone | Exponential modulated tones with 100% AM & 5% FM | 16 epochs of 1024 data points and lasted a total of 13.107 | Noninverting on Fz. Inverting on ipsilateral mastoids. | Epoch > $\pm 80$ uV in amplitude rejected. | Weighted | F ratio with $p < 0.05$ for at least 3 consecutive sweeps. | noise < 11 nV & $p > 0.05$ OR ASSR amp < 10 nV & $p \geq$ | Random start between 10 to 30 dB HL, increased or |
|  |  |  | Multiple CF: 0.5, 1, 2, & 4 kHz MF: 78, 85, 93, & 101 Hz | seconds. A min of 10 consecutive sweeps | Ground on low forehead. |  |  |  | 0.30 | decreased in 10-dB steps |
| Ismaila et al (2016) [50] | Gsi AUDERA | Mastoid | Modulated tones with 100% AM & 10% FM Multiple CF: 0.5, 1, 2, & 4 kHz MF: 74, 81, 88, & 95 Hz | NA | Noninverting on high forehead (Fz). Inverting on ipsilateral mastoid. Ground on contralateral mastoid. | | | PC<sup>2</sup> with $p < 0.03$ | 1mV | 10-dB steps |
| Valeriotte & Small (2024) [44] | MASTER II | Mastoid | Exponential modulated tones with 100% AM Multiple CF: 0.5, 1, 2, & 4 kHz MF: 78, 85, 93, & 101 Hz | 16 epochs of 1024 data points and lasted a total of 13.11 seconds. A min of 10 consecutive sweeps | Noninverting on Vertex. Inverting on ipsilateral mastoids. Ground on low forehead. | | | F ratio with $p < 0.05$ for at least 3 consecutive sweeps. | noise < 15 nV & $p > 0.05$ | Started at 30 dB HL, increased 20-dB or decreased in 10-dB steps |

To generate and record the responses, the Rotman MASTER system [51] I and II were used in eight out of 12 reports, and the GSI AUDERA (Grason-Stadler, Eden Prairie, Minnesota, USA) used twice [48,50]. The B71 bone transducer (Radioear, New Eagle, Pennsylvania, USA) was used in all reports. It was positioned on the temporal bone in nine reports, on the mastoid in two reports [48,50], and on the forehead in a report [45].

Different types of stimuli were used across reports. 100% amplitude modulated (AM) tones were used in two reports [22,45], with mixed modulated tones (100% AM and 10% to 25% FM) in another eight reports [23,24,32,46–50]. Exponentially AM tones (100% AM and 5% FM) were used two reports [33,44]. Carrier frequencies tested were 500, 1000, 2000, and 4000 Hz in all studies, with 250 Hz also tested by Swanepoel et al (2008) [48]. Online artefact rejection was set by all but two reports [48,50], with threshold for rejection of epochs between ±40 and 80 µV. Weighted [52], rather than linear averaging of sweeps was used in six reports [23,24,32,33,47,49].

#### Detection algorithm and noise criterion

Multiple detection algorithms have been employed for ASSR analysis: F ratio, Hotelling T^2^ (HT^2^), phase coherence squared (PC^2^), as well as combinations of the above [9,53]. The F ratio for hidden periodicity compares the power of the signal (i.e., the modulation frequency) to the mean power of *N* adjacent frequencies. The F ratio follows an F-distribution with [2 and *N*] degrees of freedom [7,9]. PC^2^ and HT^2^ are considered repeated measurements of the variance of the response [7], as they are based on individual epochs rather than an average epoch [9]. HT^2^ is the two-dimensional analogue of univariate t-tests, where the real and imaginary components of the epoch spectra form a two-dimensional vector. The HT^2^ test assesses whether the means of the real and imaginary components are significantly different from zero [7,9]. The PC^2^, in contrast, considers only the phase of the response and is based on the Rayleigh test [7,53].

An ASSR system requires an algorithm for determining the likelihood of the absence or presence of a response. For the former, usually a Signal to Noise Ratio (SNR) analysis and/or noise criterion threshold are involved and, for the latter, a Residual Noise (RN) analysis is required.

The efficiency with which this is carried out will determine the accuracy in threshold estimation. It has been shown that increased number of sweeps will increase SNR and reduce RN resulting in increased threshold estimation accuracy [54]. It is therefore important to consider the effects of detection technique, the effects of number of sweeps, and noise criterion for stopping averaging in the current analysis.

The detection algorithms employed depended on the device. The Rotman MASTER I and II decision is based on F ratio while the GSI AUDERA is based on PC^2^ (see [10] for a comparison). In nine reports, the noise criterion for considering a response absent was defined as either noise < 11 nV and *p* > 0.05, or noise < 10 nV and *p* ≥ 0.30 [22–24,32,33,44,46,47,49]. Other reports used a non-significant detection of ASSR for 64 sweeps [45], a noise criterion threshold of 1 µV [48] or 1 mV [50].

#### Effect measures

As stated in the pre-registration, a minimum of three papers with at least three stars on the NOS tool was required for inclusion in the synthesis.

Four reports, yielding five studies, compared BC behavioural and BC ASSR thresholds in NH adults [45,46,49,32]. The data from these studies were used to carry out a meta-analysis.

BC ASSR thresholds in NH adults were estimated in seven reports, yielding eight studies [22,24,32,33,45,47,49]. The data from these studies were used to carry out a second meta-analysis.

Thresholds in HI adults were estimated only by Ishida et al. (2011) [49], which was not sufficient to carry out a meta-analysis.

Only two studies [33,50] compared BC behavioural and BC ASSR thresholds in NH infants. These data were not used for meta-analyses.

BC ASSR thresholds in NH infants were estimated in eight reports, yielding 13 studies [23,24,32,33,46–48,50]. The data from these studies were used to carry out a third meta-analysis.

BC ASSR thresholds in infants with conductive HL at 500 Hz were estimated in three reports [44,48,50]. The data from these studies were used to carry out a fourth meta-analysis.

In total, four meta-analyses were performed: (1) BC ASSR thresholds vs BC behavioural in NH adults, (2) BC ASSR thresholds in NH adults, (3) BC ASSR thresholds in NH infants, (4) BC ASSR thresholds in conductive HL infants at 500 Hz.

#### Hearing threshold

For adult participants, behavioural AC and, in five studies, BC thresholds were obtained, so their hearing thresholds were clinically ascertained.

For infant participants, Casey & Small (2014) [33] performed visual reinforcement audiometry to estimate hearing threshold at different frequencies [55]. Ismaila et al. (2016) [50] performed play audiometry but reported only BC thresholds.

Hearing was screened via automatic auditory brain stem response (AABR) and/or distortion-product otoacoustic emissions (DPOAE) by five studies [23,24,46–48] with also tympanograms by Swanepoel et al (2008) [48]. Hearing screening in Small & Hu (2011) [31]. was based only on transient evoked otoacoustic emission (TEOAE). However, AABR and OAE do not provide frequency-specific estimates of auditory function. This is their main drawback for hearing screening [6,56]. Valeriote and Small (2024) [44] performed AC and BC ABR at multiple frequencies, tympanograms and TEOAEs, which permit more accurate screening.

#### Same participants in multiple experiments or studies

Small et al. (2007) [23] compared the effects of BC coupling methods (i.e., elastic-band vs hand-held) and oscillator placements (temporal, mastoid and forehead) on ASSR threshold. The two coupling methods were recorded in the same infants; here we included the elastic-band thresholds because their standard deviation was smaller than for the hand-held thresholds. The three oscillator placements were recorded in the same infants; here we included temporal bone thresholds as it was the placement most commonly used in the other articles.

Two reports [24,32] combined new participants’ thresholds with those previously reported, or combined thresholds from different articles. These thresholds are included here because their mean and standard deviation were different.

### Risk of bias in studies

A summary of these assessments is provided in Table 3. Six reports were classified as high risk [23,24,32,46–48] because the evaluation of hearing thresholds in infants was not frequency-specific (see above). Four reports were classified as intermediate risk [22,33,45,50], and two study as low risk [44,49]. Since none of the studies included follow-up, only one star was assigned in the Outcome category. Cumulatively, all studies received at least three stars and were therefore included in the synthesis.

**Table 3:**
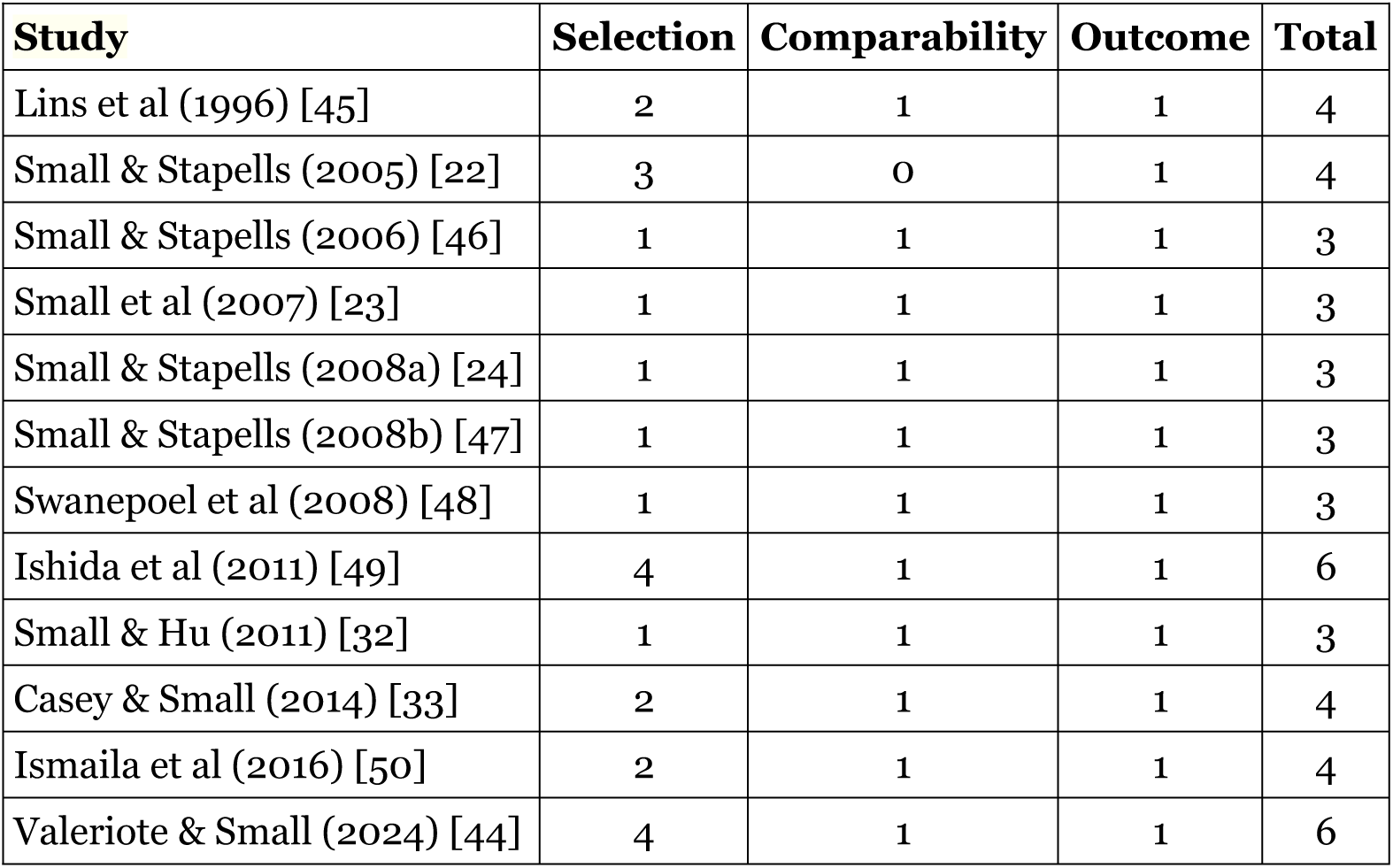
Risk of bias for each of the included study.

### Results of individual studies and syntheses

#### BC ASSR vs behavioural thresholds in NH adults

Fig 2 shows the forest plot for BC ASSR vs behavioural thresholds in NH adults as a function of the frequency (see also Supplementary Table S2). The number of participants per frequency was between 60 and 62. Heterogeneity was less than 50% only at 2000 Hz (I^2^ = 43%, τ^2^ = 6.1, *p* = 0.14). It was high at 500 (I^2^ = 52%, τ^2^ = 15.4, *p* = 0.08), 1000 (I^2^ = 68%, τ^2^ = 33.0, *p* = 0.01), and 4000 Hz (I^2^ = 58%, τ^2^ = 11.9, *p* = 0.06). For consistency, a random effects model was carried out for each frequency, using the inverse variance method, the restricted maximum-likelihood estimator for τ^2^, and untransformed (raw) means.

**Figure 2:**
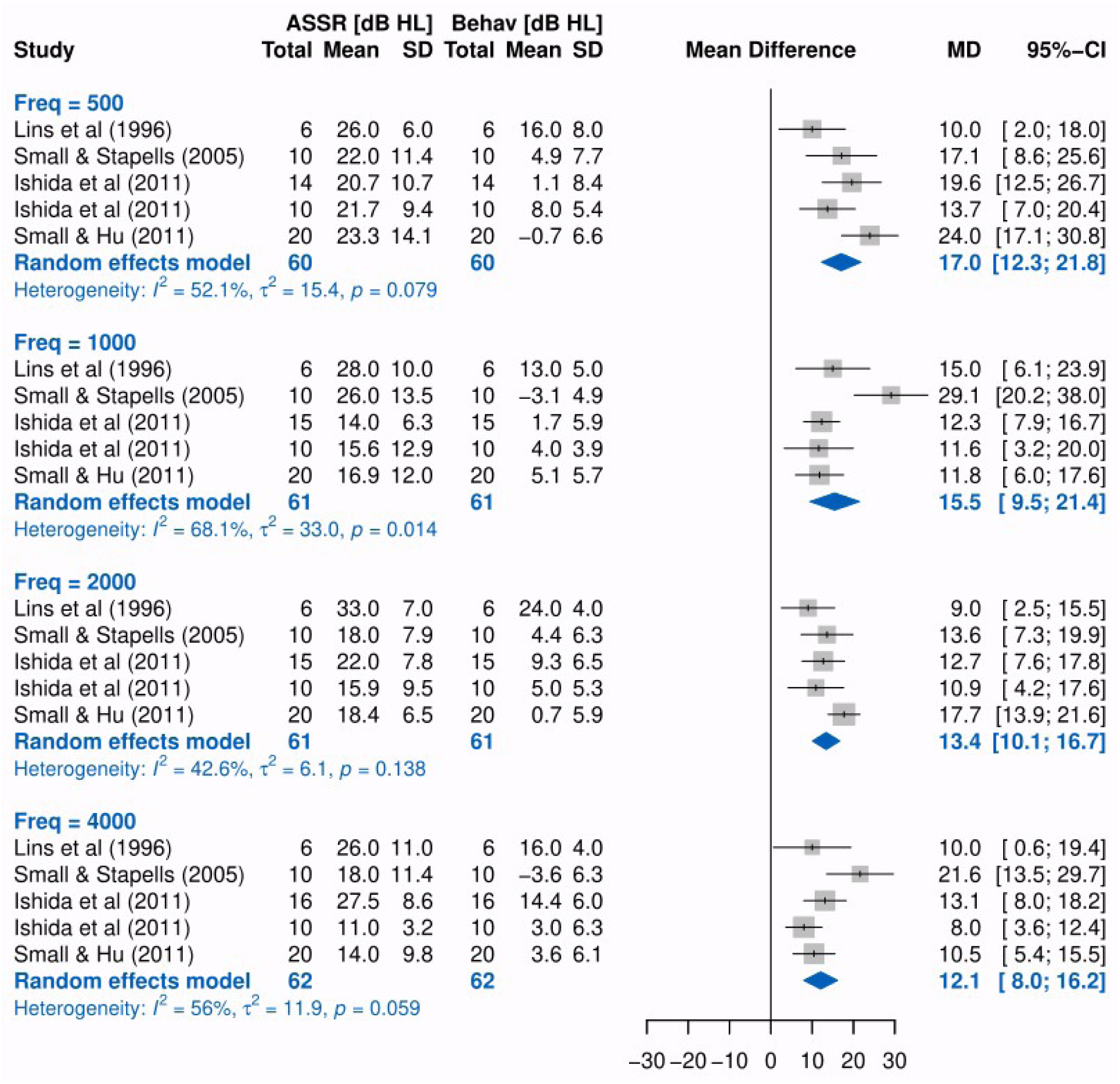
Forest plot for BC ASSR vs behavioural thresholds in NH adults as a function of the frequency. Behav: behaviour; SD: standard deviation; MD: mean difference; CI = confident intervals.

Means (and 95% confident intervals) of the random effects model for each frequency were 17.0 (12.3/21.8) dB at 500 Hz, 15.5 (9.5/21.4) dB at 1000 Hz, 13.4 (10.1/16.7) dB at 2000 Hz, and 12.1 (8.0/16.2) dB at 4000 Hz.

#### BC ASSR thresholds in NH adults

Fig 3 shows the forest plot for BC ASSR thresholds in NH adults as a function of the frequency (see also Supplementary Table S3). Seven studies reported BC ASSR thresholds in NH adults, with Ishida et al (2011) [49] reporting the thresholds on two different cohorts. The number of participants per frequency was between 97 and 99. High heterogeneity was present at each frequency as I^2^ = 62% (τ^2^ = 12.7, *p* < 0.01) at 500 Hz; I^2^ = 65% (τ^2^ = 17.9, *p* < 0.01) at 1000 Hz; I^2^ = 80% (τ^2^ = 26.3, *p* < 0.01) at 2000 Hz; and I^2^ = 89% (τ^2^ = 41.2, *p* < 0.01) at 4000 Hz.

**Figure 3:**
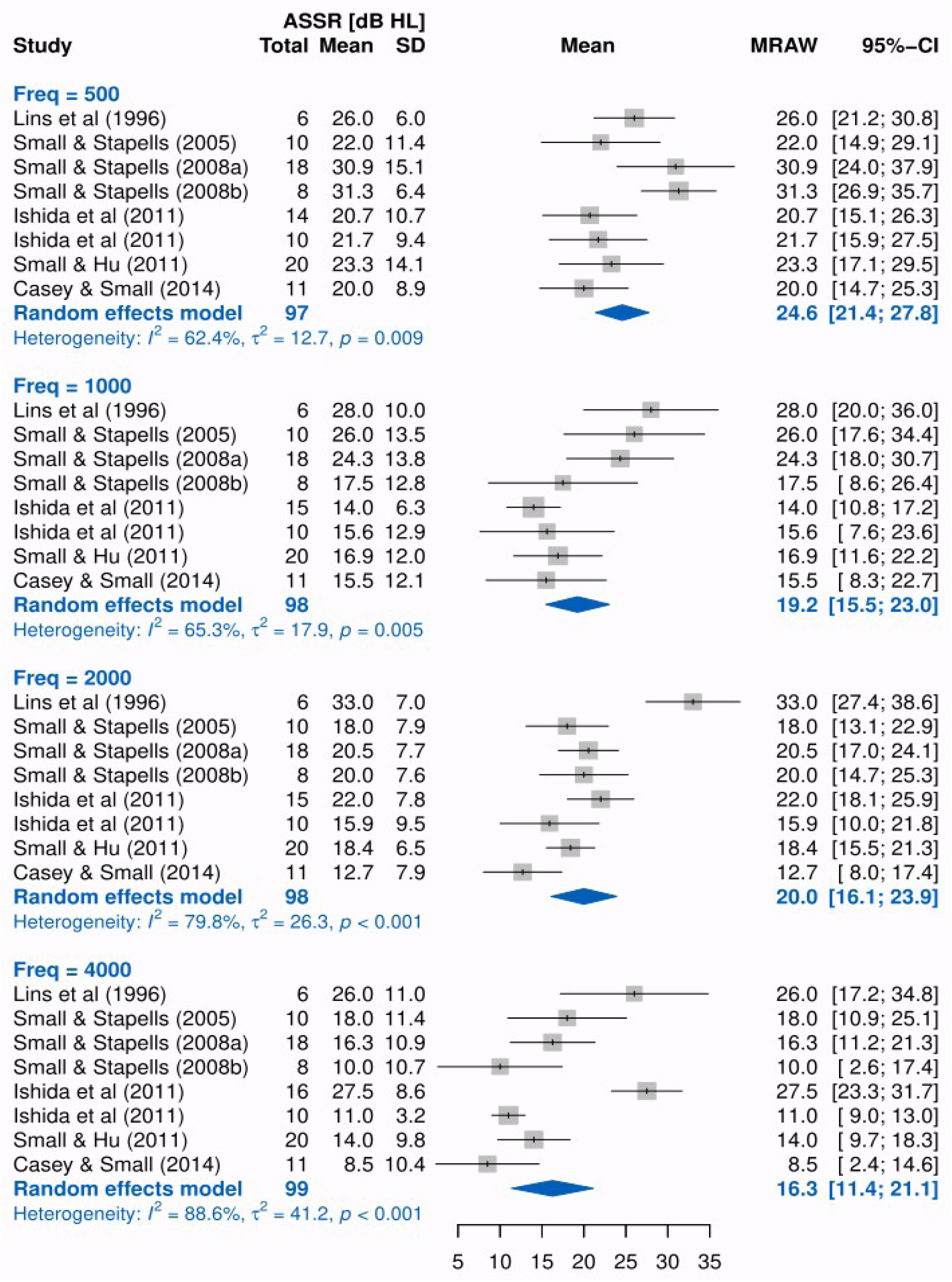
Forest plot for BC ASSR thresholds in NH adults as a function of the frequency. SD: standard deviation; MRAW: mean raw, i.e., untransformed means; CI = confident intervals.

Therefore, a random effects model was carried out, using the inverse variance method, a restricted maximum-likelihood estimator for τ^2^ and untransformed (raw) means.

Means (and 95% confident intervals) of the random effects model for each frequency were 24.6 (21.4/27.8) dB HL at 500 Hz, 19.2 (15.4/23.0) dB HL at 1000 Hz, 20.00 (16.1/23.9) dB HL at 2000 Hz, and 16.3 (11.4/21.1) dB HL at 4000 Hz.

#### BC ASSR thresholds in NH infants

Fig 4 shows the forest plot for BC ASSR thresholds in NH infants as a function of the frequency (see also Supplementary Table S4 and Table S5). Seven reports reported BC ASSR thresholds in NH infants, with additional four reports reporting the thresholds on different cohorts [23,24,32,46]. The number of participants per frequency was between 249 and 271. High heterogeneity was present at each frequency as I^2^ = 51% (τ^2^ = 7.0, *p* < 0.01) at 500 Hz; I^2^ = 93% (τ^2^ = 39.6, *p* < 0.01) at 1000 Hz; I^2^ = 81% (τ^2^ = 18.0, *p* < 0.01) at 2000 Hz; and I^2^ = 90% (τ^2^ = 48.3, *p* < 0.01) at 4000 Hz. Therefore, a random effects model was carried out, using the inverse variance method, a restricted maximum-likelihood estimator for τ^2^ and untransformed (raw) means.

**Figure 4:**
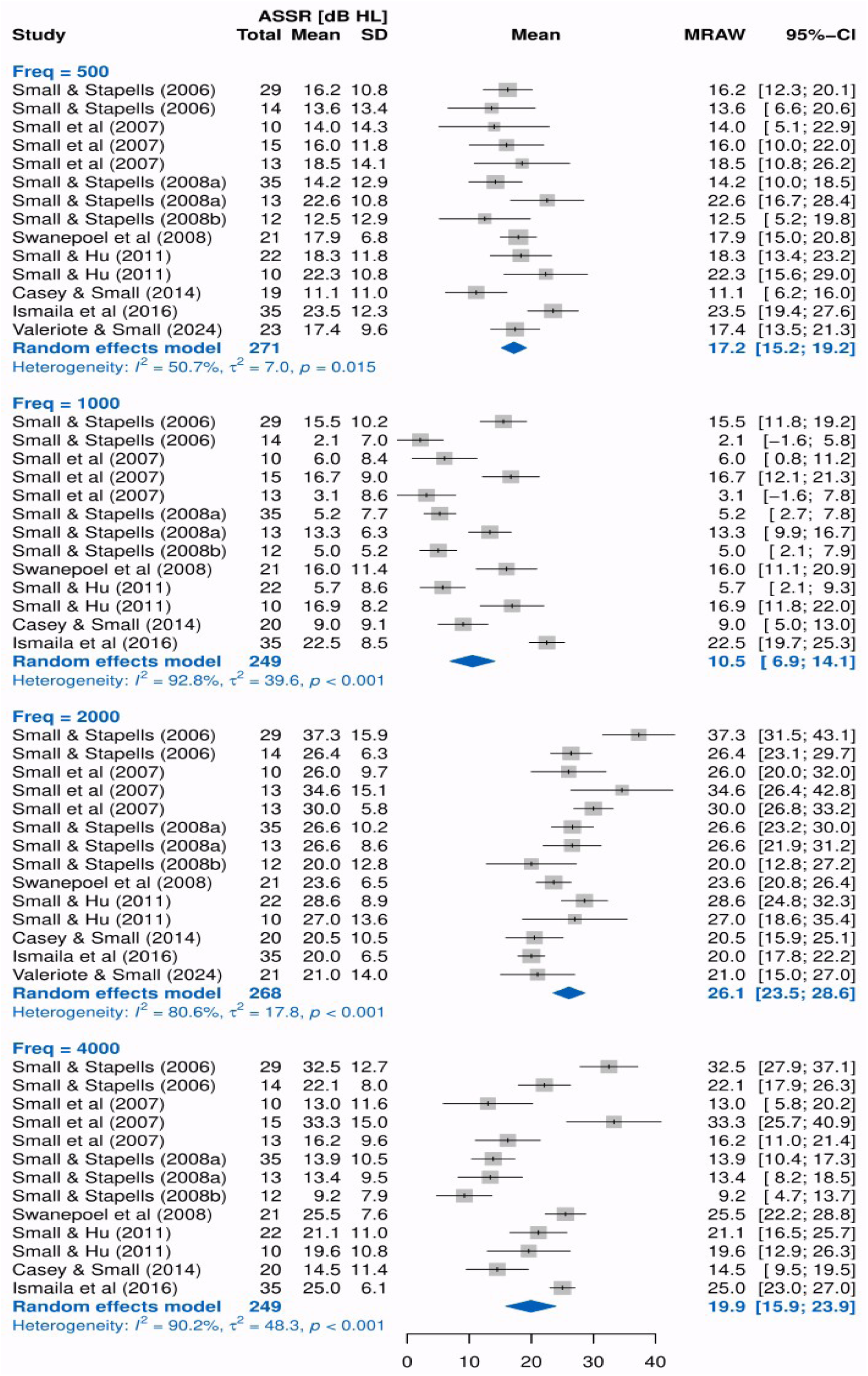
Forest plot for BC ASSR thresholds in NH infants as a function of the frequency. SD: standard deviation; MRAW: mean raw, i.e., untransformed means; CI = confident intervals.

**Figure 5:**
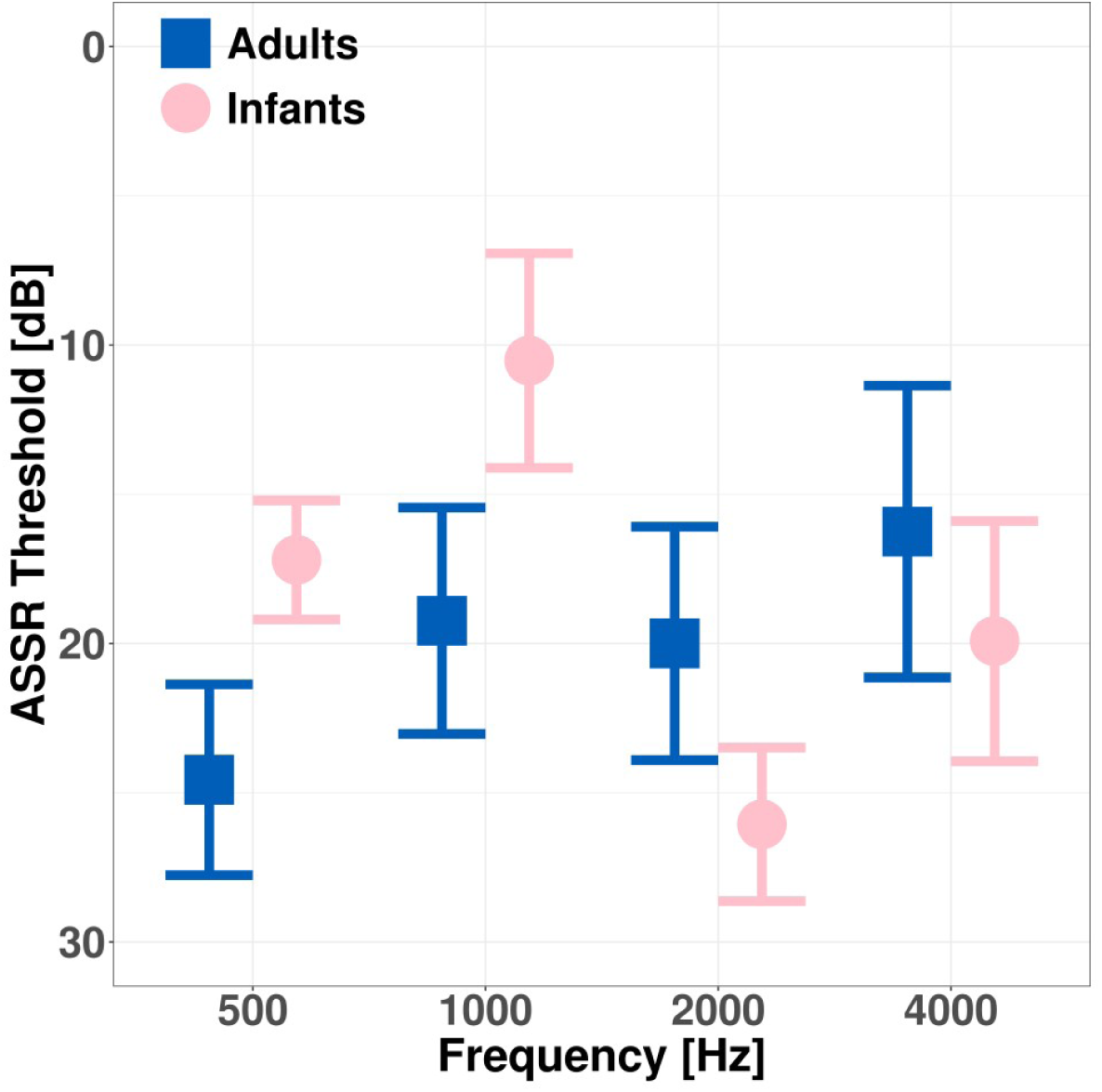
Summary of the meta-analysis results for BC ASSR thresholds in adults (beige square) and infants (purple circle). The ASSR threshold dB (attenuator) as a function of the frequency.

Means (and 95% confident intervals) of the random effects model for each frequency were 17.2 (15.2/19.2) dB HL at 500 Hz, 10.5 (6.9/14.1) dB HL at 1000 Hz, 26.1 (23.5/28.6) dB HL at 2000 Hz, and 19.9 (15.9/23.9) dB HL at 4000 Hz.

#### BC ASSR thresholds in CHL infants at 500 Hz

Three reports estimated BC ASSR thresholds in CHL infants at 500 Hz, for a total of 85 participants (see also Supplementary Table S6). High heterogeneity was present having I^2^ = 82% (τ^2^ = 12.9, *p* < 0.01). Therefore, a random effects model was carried out, using the inverse variance method, a restricted maximum-likelihood estimator for τ^2^ and untransformed (raw) means. Mean (and 95% confident intervals) of the random effects model was 20.3 (15.6/24.9) dB HL.

#### Spurious responses

Of particular interest are the minimum intensity levels at which spurious (artefactual) results were observed. Table 4 summarises the spurious results reported in the included studies. The level at which spurious responses occur increased as a function of frequency. Similarly, the percentage of participants with spurious responses increased with frequency. Therefore, the BC ASSR threshold at low frequencies, such as 500 Hz, and moderate intensities, such as 40 dB HL, may not be reliable. Swanepoel and colleagues [48] questioned the clinical value of BC ASSR thresholds measured at or below 500 Hz. Current UK guidance recommends that BC ASSR should not be performed at 500 Hz and beyond 40 dB nHL for frequencies between 1000 and 4000 Hz [12].

**Table 4:** Spurious (non-auditory) responses reported in the included papers. SNHL: sensorineural hearing loss.

| Study | Note |
| --- | --- |
| Small & Stapells (2005) [22] | The percentage of participants who had thresholds $\leq 20$ dB HL were 60, 40, 90, and 60% at 500, 1000, 2000, and 4000 Hz, respectively.<br>The percentage of participants who had thresholds $\leq 30$ dB HL were 90, |
|  | 70, 100, and 100% at 500, 1000, 2000, and 4000 Hz, respectively. |
| Small & Stapells (2006) [46] | For the post-term infants, 90% or more of the subjects had responses present at minimum intensities of 30, 10, 40, and 30 dB HL for 500, 1000, 2000, and 4000 Hz, respectively.<br>For the preterm infants, 90% or more of the subjects had responses present at minimum intensities of 40, 30, 50, and 50 dB HL for 500, 1000, 2000, and 4000 Hz, respectively. |
| Swanepoel et al (2008) [48] | Spurious BC responses for children with severe-to-profound SNHL. See their Table 4 for % spurious for children with mild-to-moderate SNHL. Min levels where spurious results were seen: 40 dB HL @ 500 Hz; 60 dB HL @ 1000, 2000, 4000. |
| Ismaila et al (2016) [50] | Min levels where spurious results were seen: 52 dB HL @ 500 Hz; 66.5 dB HL @ 1000 Hz; 69 dB HL @ 2000 Hz; 64 dB HL @ 4000 Hz. |

BC stimuli presented at moderate and high intensities elicited spurious responses, primarily at 500 Hz and 1000 Hz. At these frequencies and range of intensities, BC ASSR amplitudes were abnormally larger when compared either to those at low intensities or to AC ASSR amplitudes [57]. The reasons for these findings, along with potential solutions, are discussed elsewhere [22,57,58].

### Certainty assessment

The GRADE Summary of Findings tables for BC ASSR thresholds versus behavioural thresholds in NH adults are presented in Table 5, those for BC ASSR thresholds in NH adults and infants are presented in Table 6, and those for BC ASSR in CHL infant in Table 7. As the included studies were cohort studies, the body of evidence began with a low-certainty rating. The certainty was subsequently downgraded to very low due to serious inconsistency, reflected by high heterogeneity (in both adults and infants), and serious indirectness (in infants), as the evaluation of hearing thresholds was not frequency-specific.

**Table 5:**
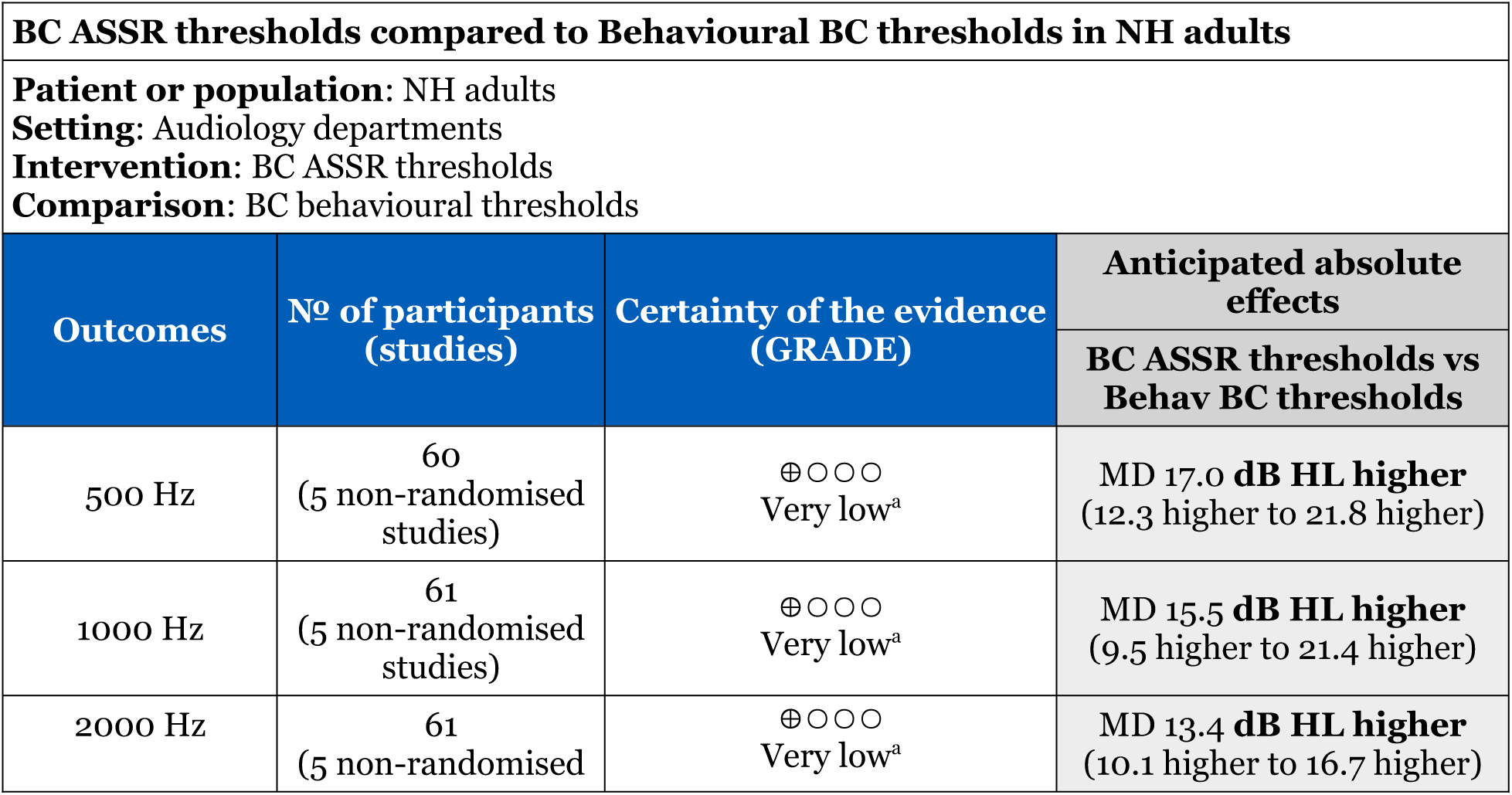
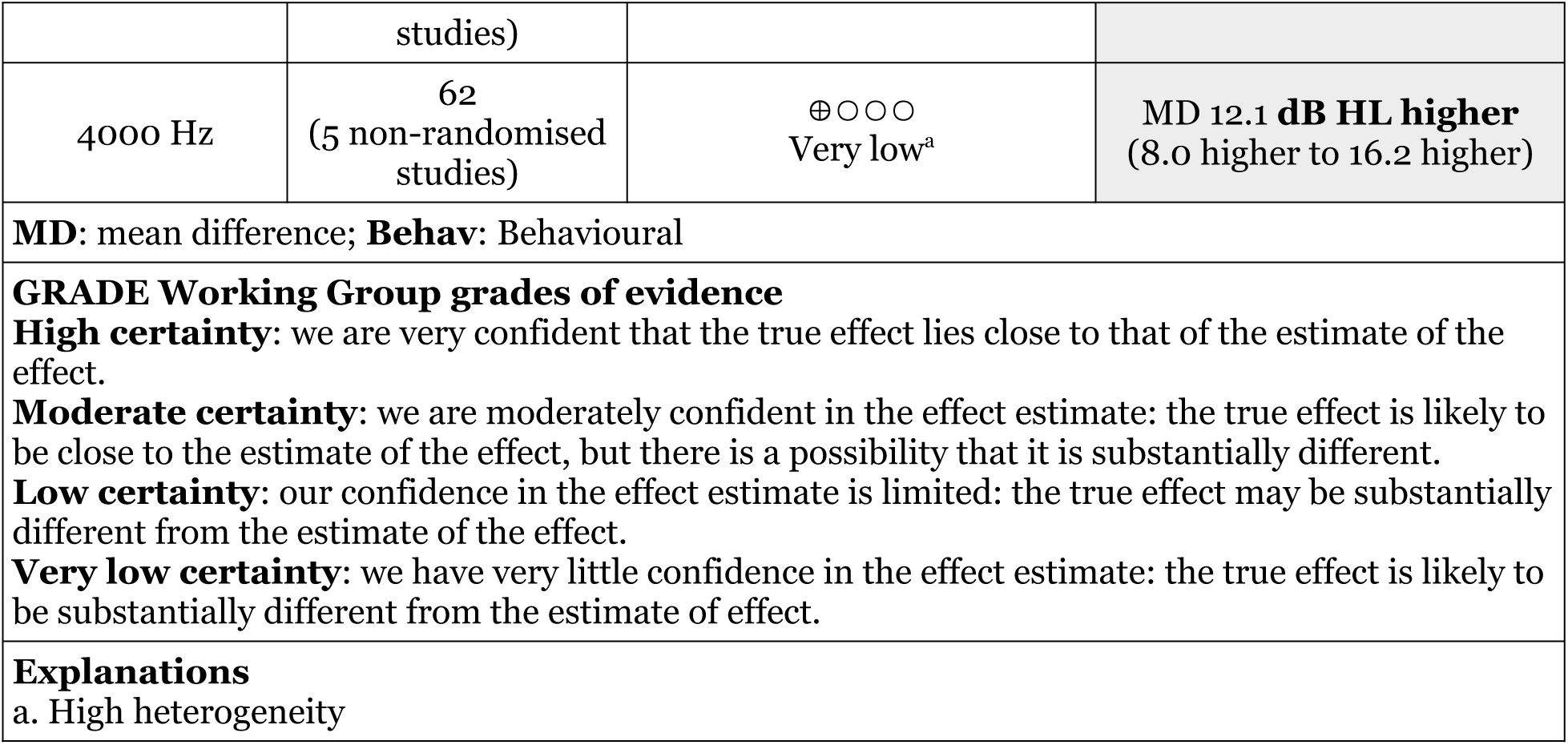
Summary of Findings tables for BC ASSR thresholds versus behavioural thresholds in NH adults.

**Table 6:**
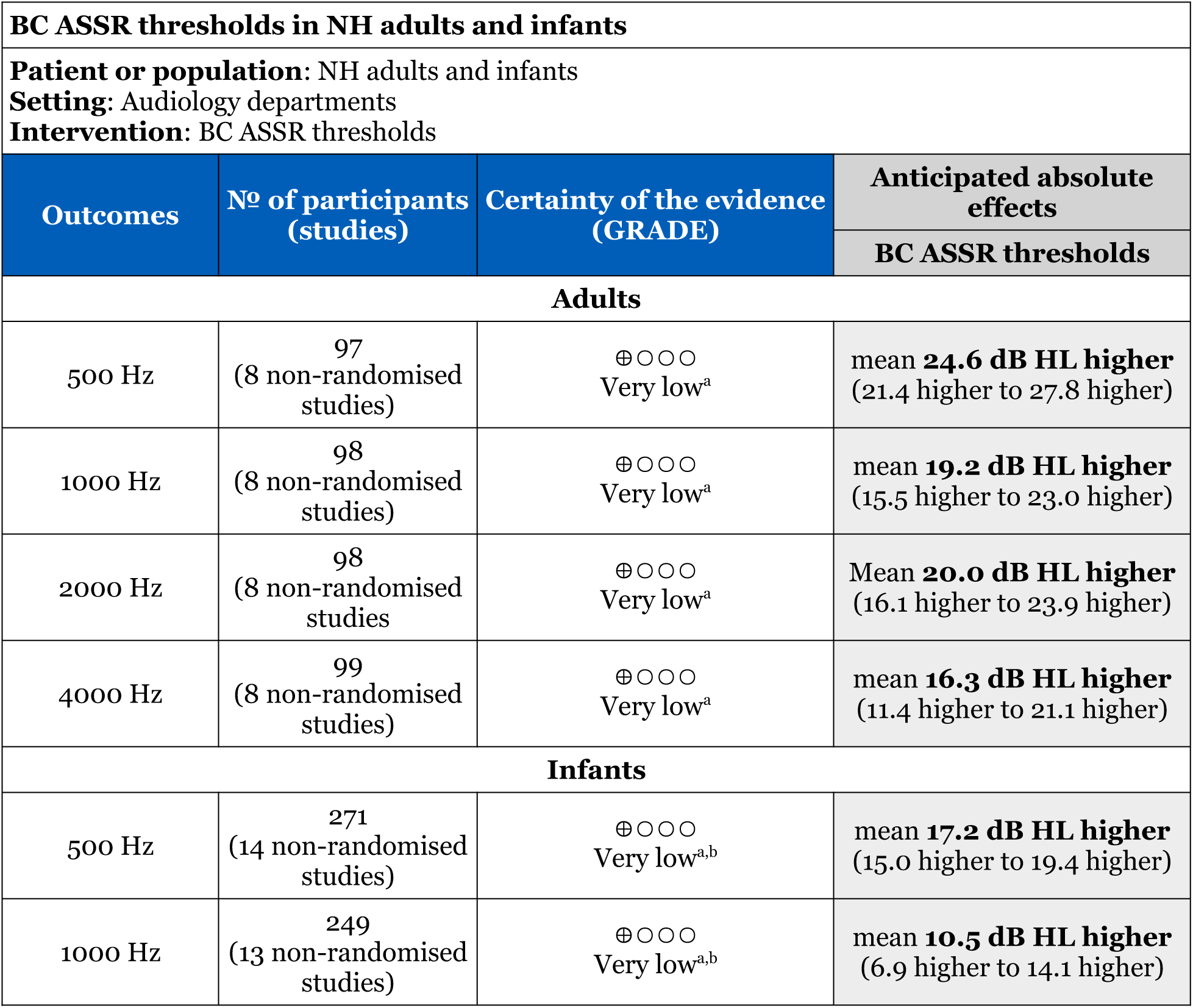
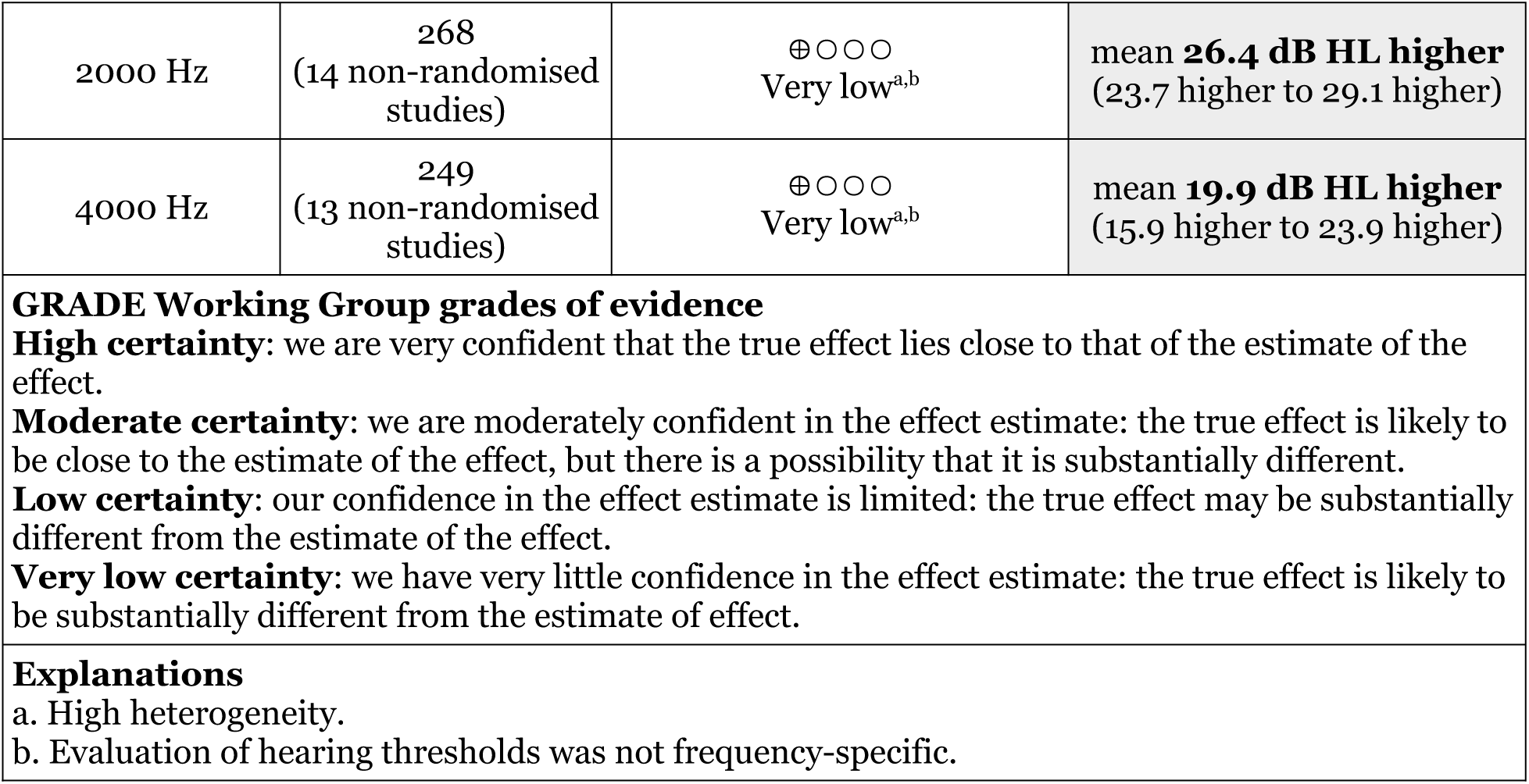
Summary of Findings tables for BC ASSR thresholds in NH adults and infants.

## Discussion

The present meta-analysis included 11 papers spanning 28 years of research in BC ASSR. No other available meta-analyses have specifically considered BC ASSR or BC ABR.

An accurate BC ASSR threshold estimation relies on precise and reliable “correction” values that allow the prediction of behavioural thresholds from the measured ASSRs values. A possible definition of a reliable BC ASSR threshold is that its 95% confidence interval is within 10 dB of the mean of each model. The width of the confidence interval ranged from 2.2 to 6.0 dB HL, indicating that it remained within 10 dB.

Our main finding is the elevation of BC ASSR thresholds compared to behavioural thresholds, as a function of both age group and frequency. For NH adults, the difference between behavioural and ASSR thresholds ranged from 12.1 to 17.0 dB, while ASSR thresholds ranged from 16.3 to 24.6 dB HL across the frequency range of 500-4000 Hz. For NH infants, ASSR thresholds ranged from 10.5 to 26.4 dB HL within the same frequency range.

The BSA Early Assessment Guidance [[59], Appendix H] provides provisional corrections for ABR thresholds as a function of frequency and age. For NH adults, the provisional corrections range from 11.8 to 20.4 dB, which are similar to our findings. However, for NH infants, the provisional corrections range from 13.6 to 19.6 dB, which differ from our findings. Therefore, new correction values should account for the elevations reported here.

To explore a less-well established technique such as BC ASSRs, thresholds should be compared with a gold standard technique such as behavioural thresholds. This is achievable in adults, but not possible in neonates as behavioural assessment only becomes possible around seven months and even then, a lengthy period may be required to determine accurate behavioural thresholds. A compromise would be to compare the BC ASSR thresholds to those obtained using BC ABR.

Our original interest was to run meta-analyses on BC ASSR thresholds in HI adults and infants. However, only three studies reported BC ASSR thresholds for these groups: one study for HI adults and two for HI infants. Therefore, only NH thresholds were analysed.

### Heterogeneity

Heterogeneity was strongly apparent across the studies. This was due to multiple factors, such as stimuli, statistical indices, stopping criteria, and threshold definitions used. Furthermore, several studies reported a large standard deviation (up to 16.9 dB HL) of BC ASSR thresholds. Nevertheless, the average BC ASSR thresholds were consistently positive, meaning they were always higher than the behavioural thresholds.

Because of the small number of studies available, it was not possible to assess the effect of recording parameters on the BC ASSR thresholds. A meta-analysis on AC ASSR thresholds revealed that the effect of recording parameters, such as type of modulation (i.e., amplitude or mixed modulated) and maximum number of sweeps, varied for different frequencies and hearing status [19].

| BC ASSR thresholds in CHL infants |  |  |  |
| --- | --- | --- | --- |
| **Patient or population:** CHL infants |  |  |  |
| **Setting:** Audiology departments |  |  |  |
| **Intervention:** BC ASSR thresholds |  |  |  |
| Outcomes | Nº of participants (studies) | Certainty of the evidence (GRADE) | Anticipated absolute effects |
|  |  |  | BC ASSR thresholds vs Behav BC thresholds |
| 500 Hz | 85 | ⊕○○○ | mean 20.3 dB HL higher |
|  | (3 non-randomised studies) | Very low<sup>a,b</sup> | (15.6 higher to 24.9 higher) |
| **GRADE Working Group grades of evidence**<br>**High certainty:** we are very confident that the true effect lies close to that of the estimate of the effect.<br>**Moderate certainty:** we are moderately confident in the effect estimate: the true effect is likely to be close to the estimate of the effect, but there is a possibility that it is substantially different.<br>**Low certainty:** our confidence in the effect estimate is limited: the true effect may be substantially different from the estimate of the effect.<br>**Very low certainty:** we have very little confidence in the effect estimate: the true effect is likely to be substantially different from the estimate of effect. |  |  |  |
| **Explanations**<br>a. High heterogeneity<br>b. Evaluation of hearing thresholds was not frequency-specific. |  |  |  |

### Infant thresholds

The measure of BC ASSR thresholds for infants and children should be interpreted with extra caution because of their age. Specifically, the reported age varied from 34.5 weeks PCA to 4.5 years. Including such a large age range might be a confound for BC ASSR thresholds as, at least for AC ASSR thresholds, thresholds tend to decrease in NH infants as a function of age [60,61]. Our rationale for including the full age range was that the heterogeneity and variability of the studies were greater than any potential effects of age. For instance, ASSR threshold (and standard deviation) at 2000 Hz was nearly independent of Age: 20.0 (12.8) dB HL in 21-week-old infants [47], 20.50 (10.5) dB HL in 10.5-month infants [33], 23.6 (6.5) dB HL in 3.6-year-old children [48], and 20.0 (6.5) dB HL in 4.5-year-old children [50]. However, to rule out the possible confound due to Age, a further meta-analysis was conducted excluding the studies in children [48,50]. Means (and 95% confident intervals) of the random effects model for each frequency were 16.4 (14.5/18.3) dB HL at 500 Hz, 8.9 (5.6/12.1) dB HL at 1000 Hz, 27.0 (24.3/29.7) dB HL at 2000 Hz, and 18.9 (14.3/23.4) dB HL at 4000 Hz. Thus, the overall mean difference was 0.65 dB HL between the model including children and the one excluding them.

### BC ASSR thresholds for adults and infants

Among others, Casey & Small (2014) [33] compared BC ASSR thresholds in adults and infants. Fig 6 shows a summary of the meta-analysis results for BC ASSR thresholds in adults (blue squares) and infants (pink circles) in the same conditions, i.e., only ASSR.

Adult thresholds are higher at low frequencies (500 and 1000 Hz) and lower at high frequencies (2000 and 4000 Hz) when compared to those of infants. The adult threshold at 2000 Hz was about the same of those at 1000 and 4000 Hz. Instead, the infant threshold at 2000 Hz was clearly higher than those at other frequencies. Since BC ASSR thresholds change as a function of age and frequency, correction factors are needed for different population and frequency.

### Variability of BC ASSR thresholds

In the current meta-analyses, the standard deviations for BC ASSR thresholds across frequency was between 16.1 (at 500 Hz) and 24.8 (at 4000 Hz) dB HL for adults, and between 16.3 (at 500 Hz) and 32.4 (at 4000 Hz) dB HL for infants. Comparing these with AC ASSR thresholds, the standard deviation was reported between 9.9 and 12.1 dB HL for adults [19], and between 7 and 15 dB SL for infants (Table 9-2 in [13]). Comparing BC ASSR with BC tone burst ABR, the standard deviation was between 6.2 and 10.5 dB HL for adults, and between 7.2 and 10 for infants [62,63]. Therefore, the inter-subject variability across frequency in BC ASSR threshold was larger than those in AC ASSR and BC ABR thresholds.

It has been suggested that the substantial variability in BC ASSR thresholds may be due to factors such as variations in transducer placement, the condition of occluded versus non-occluded ears, differences in stimulus calibration methods [22]. Other possible contributing factors include test duration, artefact rejection thresholds, average type, detection algorithms, and noise criteria.

The aforementioned standard deviation values refer to inter-subject variability. Since BC ASSR thresholds can be influenced by several factors, intra-subject variability should also be evaluated. Future studies should assess the test-retest reliability of BC ASSR threshold [64].

### Possible limitations

Beside the range of age of the infants discussed earlier, another limitation was that most of the included studies were conducted by Stapells and Small. Contribution from other laboratories or countries should help to improve the quality of research such as, for instance, removing risk of bias. A related issue was that most of the thresholds were collected using the Rotman MASTER system, a first-generation system that is no longer available. The lack of thresholds obtained using a second generation system [20] should be addressed in future research.

There were also limitations in the review processes used. For instance, the exclusion of non-English articles and conference abstracts, which could have improved the quality of the research by diversifying the data. Nevertheless, we are confident in our outcome and the overall conclusions.

### Conclusions

Findings from this meta-analysis suggest that ASSR can be a reliable technique to estimate BC thresholds in NH populations. In the frequency range of 500-4000 Hz, the ASSR threshold ranged from 16 to 24 dB HL in NH adults, and from 10 to 26 dB HL in NH infants. Conductive HL leads to an elevation of 20 dB HL at 500 Hz. Future work is needed to expand the measure of BC ASSR thresholds in NH infants, as well as to start investigating both HI adults and infants. Only then can we reliably define normative BC ASSR thresholds for different populations.

## Declarations

### Registration

This systematic review has been registered in the international prospective register of systematic reviews (PROSPERO) under the registration number: CRD42023422150.

## Author contributions

EP: Conceptualisation, Methodology, Software, Validation, Formal analysis, Investigation, Resources, Data Curation, Writing - Original Draft, Writing - Review & Editing, Visualization, Supervision, Project administration. CG: Conceptualisation, Methodology, Validation, Investigation, Resources, Data Curation, Writing - Original Draft, Writing - Review & Editing, Supervision, Project administration.

## Availability of data and material

All data generated or analyses during this study are included in this published article and its Supplementary Table S5.

## Competing interests

The authors declare that they have no competing interests.

## Funding

E.P. is supported by the NIHR Manchester Biomedical Research Centre (NIHR203308).

## Supporting information

PRISMA 2020 checklist

## Data Availability

All data generated or analyses during this study are included in this published article and its Supplementary Table S7.

## Acknowledgments

We thank Timothy S. Wilding and Garreth Prendergast for helpful comments on an earlier version of this paper.

## Supporting information

**S1 Table**. Search strategy

**S2 Table**. BC ASSR vs behavioural thresholds in NH adults as a function of the frequency.

**S3 Table**. BC ASSR thresholds in NH adults as a function of the frequency.

**S4 Table**. BC ASSR thresholds in NH infants at 500 and 100 Hz.

**S5 Table**. BC ASSR thresholds in NH infants at 2000 and 4000 Hz.

**S6 Table**. BC ASSR thresholds in CHL infants at 500 Hz.

**S7 Table**. Data.

## Notes

### Competing Interest Statement

The authors have declared no competing interest.

### Author Declarations

This study was a meta-analysis based on data extracted from previous studies.

### Summary of Updates

The meta-analysis was updated to include a new report.

