## Supplementary tables for "A meta-analysis of bone conduction 80 Hz auditory steady state response thresholds in adults and infants"

| Database | Search details |
|---|---|
| PubMed | ("Bone Conduction" OR "Bone-Conducted" OR "Bone Vibrator")<br>AND<br>("ASSR" OR "Auditory Steady-State Response" OR<br>"EFR" OR "Envelope Following Response") |
| Cochrane Library | #1 MeSH descriptor: [Bone Conduction] explode all trees<br>#2 (Bone Conduction):ti,ab,kw<br>#3 (Bone-Conducted):ti,ab,kw<br>#4 (Bone Vibrator):ti,ab,kw<br>#5 #1 OR #2 OR #3 OR #4<br>#6 (Auditory Steady-State Response):ti,ab,kw<br>#7 (ASSR):ti,ab,kw<br>#8 (Envelope Following Response):ti,ab,kw<br>#9 (EFR):ti,ab,kw<br>#10 #6 OR #7 OR #8 OR #9<br>#11 #5 AND #10 |
| Embase | (("Bone Conduction" OR "Bone-Conducted" OR "Bone Vibrator")<br>AND ("ASSR" OR "Auditory Steady-State Response" OR "EFR"<br>OR "Envelope Following Response")).mp. [mp=title, abstract,<br>heading word, drug trade name, original title, device manufacturer,<br>drug manufacturer, device trade name, keyword heading word,<br>floating subheading word, candidate term word] |

**Table S1:** Search strategy

| Study | ASSR [dB HL] Total | Mean | SD | Behavioural [dB HL] Total | Mean | SD | MD | 95%-CI |
|---|---|---|---|---|---|---|---|---|
| **Freq = 500 Hz** | | | | | | | | |
| Lins et al. (1996) | 6 | 26.0 | 6.0 | 6 | 16.0 | 8.0 | 10.0 | [2.0; 18.0] |
| Small and Stapells (2005) | 10 | 22.0 | 11.4 | 10 | 4.9 | 7.7 | 17.1 | [8.6; 25.6] |
| Ishida et al. (2011) | 14 | 20.7 | 10.7 | 14 | 1.1 | 8.4 | 19.6 | [12.5; 26.7] |
| Ishida et al. (2011) | 10 | 21.7 | 9.4 | 10 | 8.0 | 5.4 | 13.7 | [7.0; 20.4] |
| Small and Hu (2011) | 20 | 23.3 | 14.1 | 20 | -0.7 | 6.6 | 24.0 | [17.1; 30.8] |
| **Random Effect model** | **60** | | | **60** | | | 17.0 | **[12.3; 21.8]** |
| **Freq = 1000 Hz** | | | | | | | | |
| Lins et al. (1996) | 6 | 28.0 | 10.0 | 6 | 13.0 | 5.0 | 15.0 | [6.1; 23.9 ] |
| Small and Stapells (2005) | 10 | 26.0 | 13.5 | 10 | -3.1 | 4.9 | 29.1 | [20.2; 38.0] |
| Ishida et al. (2011) | 15 | 14.0 | 6.3 | 15 | 1.7 | 5.9 | 12.3 | [7.9; 16.7] |
| Ishida et al. (2011) | 10 | 15.6 | 12.9 | 10 | 4.0 | 3.9 | 11.6 | [3.2; 20.0] |
| Small and Hu (2011) | 20 | 16.9 | 12.0 | 20 | 5.1 | 5.7 | 11.8 | [6.0; 17.6] |
| **Random Effect model** | **61** | | | **61** | | | 15.5 | **[9.5; 21.4]** |
| **Freq = 2000 Hz** | | | | | | | | |
| Lins et al. (1996) | 6 | 33.0 | 7.0 | 6 | 24.0 | 4.0 | 9.0 | [2.5; 15.5] |
| Small and Stapells (2005) | 10 | 18.0 | 7.9 | 10 | 4.4 | 6.3 | 13.6 | [7.3; 19.9] |
| Ishida et al. (2011) | 15 | 22.0 | 7.8 | 15 | 9.3 | 6.5 | 12.7 | [7.6; 17.8] |
| Ishida et al. (2011) | 10 | 15.9 | 9.5 | 10 | 5.0 | 5.3 | 10.9 | [4.2; 17.6] |
| Small and Hu (2011) | 20 | 18.4 | 6.5 | 20 | 0.7 | 5.9 | 17.7 | [13.9; 21.6] |
| **Random Effect model** | **61** | | | **61** | | | 13.4 | **[10.1; 16.7]** |
| **Freq = 4000 Hz** | | | | | | | | |
| Lins et al. (1996) | 6 | 26.0 | 11.0 | 6 | 16.0 | 4.0 | 10.0 | [0.6; 19.4] |
| Small and Stapells (2005) | 10 | 18.0 | 11.4 | 10 | -3.6 | 6.3 | 21.6 | [13.5; 29.7] |
| Ishida et al. (2011) | 16 | 27.5 | 8.6 | 16 | 14.4 | 6.0 | 13.1 | [8.0; 18.2] |
| Ishida et al. (2011) | 10 | 11.0 | 3.2 | 10 | 3.0 | 6.3 | 8.0 | [3.6; 12.4] |
| Small and Hu (2011) | 20 | 14.0 | 9.8 | 20 | 3.6 | 6.1 | 10.5 | [5.4; 15.5] |
| **Random Effect model** | **62** | | | **62** | | | 12.1 | **[8.0; 16.2]** |

**Table S2:** BC ASSR vs behavioural thresholds in NH adults as a function of the frequency.

| Study | Total | Mean | SD | MRAW | 95%-CI |
|---|---|---|---|---|---|
| **Freq = 500 Hz** | | | | | |
| Lins et al. (1996) | 6 | 26.0 | 6.0 | 26.0 | [21.2; 30.8] |
| Small and Stapells (2005) | 10 | 22.0 | 11.4 | 22.0 | [14.9; 29.1] |
| Small and Stapells (2008a) | 18 | 30.9 | 15.1 | 30.9 | [24.0; 37.9] |
| Small and Stapells (2008b) | 8 | 31.3 | 6.4 | 31.3 | [26.9; 35.7] |
| Ishida et al. (2011) | 14 | 20.7 | 10.7 | 20.7 | [15.1; 26.3] |
| Ishida et al. (2011) | 10 | 21.7 | 9.4 | 21.7 | [15.9; 27.5] |
| Small and Hu (2011) | 20 | 23.3 | 14.1 | 23.3 | [17.1; 29.5] |
| Casey and Small (2014) | 11 | 20.0 | 8.9 | 20.0 | [14.7; 25.3] |
| **Random Effect model** | **97** | | | **24.6** | **[21.4; 27.8]** |
| **Freq = 1000 Hz** | | | | | |
| Lins et al. (1996) | 6 | 28.0 | 10.0 | 28.0 | [20.0; 36.0] |
| Small and Stapells (2005) | 10 | 26.0 | 13.5 | 26.0 | [17.6; 34.4] |
| Small and Stapells (2008a) | 18 | 24.3 | 13.8 | 24.3 | [18.0; 30.7] |
| Small and Stapells (2008b) | 8 | 17.5 | 12.8 | 17.5 | [8.6; 26.4] |
| Ishida et al. (2011) | 15 | 14.0 | 6.3 | 14.0 | [10.8; 17.2] |
| Ishida et al. (2011) | 10 | 15.6 | 12.9 | 15.6 | [7.6; 23.6] |
| Small and Hu (2011) | 20 | 16.9 | 12.0 | 16.9 | [11.6; 22.2] |
| Casey and Small (2014) | 11 | 15.5 | 12.1 | 15.5 | [8.3; 22.7] |
| **Random Effect model** | **98** | | | **19.2** | **[15.5; 23.0]** |
| **Freq = 2000 Hz** | | | | | |
| Lins et al. (1996) | 6 | 33.0 | 7.0 | 33.0 | [27.4; 38.6] |
| Small and Stapells (2005) | 10 | 18.0 | 7.9 | 18.0 | [13.1; 22.9] |
| Small and Stapells (2008a) | 18 | 20.5 | 7.7 | 20.5 | [17.0; 24.1] |
| Small and Stapells (2008b) | 8 | 20.0 | 7.6 | 20.0 | [14.7; 25.3] |
| Ishida et al. (2011) | 15 | 22.0 | 7.8 | 22.0 | [18.1; 25.9] |
| Ishida et al. (2011) | 10 | 15.9 | 9.5 | 15.9 | [10.0; 21.8] |
| Small and Hu (2011) | 20 | 18.4 | 6.5 | 18.4 | [15.5; 21.3] |
| Casey and Small (2014) | 11 | 12.7 | 7.9 | 12.7 | [8.0; 17.4] |
| **Random Effect model** | **98** | | | **20.0** | **[16.1; 23.9]** |
| **Freq = 4000 Hz** | | | | | |
| Lins et al. (1996) | 6 | 26.0 | 11.0 | 26.0 | [17.2; 34.8] |
| Small and Stapells (2005) | 10 | 18.0 | 11.4 | 18.0 | [10.9; 25.1] |
| Small and Stapells (2008a) | 18 | 16.3 | 10.9 | 16.3 | [11.2; 21.3] |
| Small and Stapells (2008b) | 8 | 10.0 | 10.7 | 10.0 | [2.6; 17.4] |
| Ishida et al. (2011) | 16 | 27.5 | 8.6 | 27.5 | [23.3; 31.7] |
| Ishida et al. (2011) | 10 | 11.0 | 3.2 | 11.0 | [9.0; 13.0] |
| Small and Hu (2011) | 20 | 14.0 | 9.8 | 14.0 | [9.7; 18.3] |
| Casey and Small (2014) | 11 | 8.5 | 10.4 | 8.5 | [2.4; 14.6] |
| **Random Effect model** | **99** | | | **16.3** | **[11.4; 21.1]** |

**Table S3:** BC ASSR thresholds in NH adults as a function of the frequency.

| Study | Total | Mean | SD | MRAW | 95%-CI |
|---|---|---|---|---|---|
| **Freq = 500 Hz** | | | | | |
| Small and Stapells (2006) | 29 | 16.2 | 10.8 | 16.2 | [12.3; 20.1] |
| Small and Stapells (2006) | 14 | 13.6 | 13.4 | 13.6 | [6.6; 20.6] |
| Small et al. (2007) | 10 | 14.0 | 14.3 | 14.0 | [5.1; 22.9] |
| Small et al. (2007) | 15 | 16.0 | 11.8 | 16.0 | [10.0; 22.0] |
| Small et al. (2007) | 13 | 18.5 | 14.1 | 18.5 | [10.8; 26.2] |
| Small and Stapells (2008a) | 35 | 14.2 | 12.9 | 14.2 | [10.0; 18.5] |
| Small and Stapells (2008a) | 13 | 22.6 | 10.8 | 22.6 | [16.7; 28.4] |
| Small and Stapells (2008b) | 12 | 12.5 | 12.9 | 12.5 | [5.2; 19.8] |
| Swanepoel et al. (2008) | 21 | 17.9 | 6.8 | 17.9 | [15.0; 20.8] |
| Small and Hu (2011) | 22 | 18.3 | 11.8 | 18.3 | [13.4; 23.2] |
| Small and Hu (2011) | 10 | 22.3 | 10.8 | 22.3 | [15.6; 29.0] |
| Casey and Small (2014) | 19 | 11.1 | 11.0 | 11.1 | [6.2; 16.0] |
| Ismaila et al. (2016) | 35 | 23.5 | 12.3 | 23.5 | [19.4; 27.6] |
| Valeriote and Small (2024) | 23 | 17.4 | 9.6 | 17.4 | [13.5; 21.3] |
| **Random Effect model** | **271** | | | **17.2** | **[15.2; 19.2]** |
| **Freq = 1000 Hz** | | | | | |
| Small and Stapells (2006) | 29 | 15.5 | 10.2 | 15.5 | [11.8; 19.2] |
| Small and Stapells (2006) | 14 | 2.1 | 7.0 | 2.1 | [-1.6; 5.8] |
| Small et al. (2007) | 10 | 6.0 | 8.4 | 6.0 | [0.8; 11.2] |
| Small et al. (2007) | 15 | 16.7 | 9.0 | 16.7 | [12.1; 21.3] |
| Small et al. (2007) | 13 | 3.1 | 8.6 | 3.1 | [-1.6; 7.8] |
| Small and Stapells (2008a) | 35 | 5.2 | 7.7 | 5.2 | [2.7; 7.8] |
| Small and Stapells (2008a) | 13 | 13.3 | 6.3 | 13.3 | [9.9; 16.7] |
| Small and Stapells (2008b) | 12 | 5.0 | 5.2 | 5.0 | [2.1; 7.9 ] |
| Swanepoel et al. (2008) | 21 | 16.0 | 11.4 | 16.0 | [11.1; 20.9] |
| Small and Hu (2011) | 22 | 5.7 | 8.6 | 5.7 | [2.1; 9.3] |
| Small and Hu (2011) | 10 | 16.9 | 8.2 | 16.9 | [11.8; 22.0] |
| Casey and Small (2014) | 20 | 9.0 | 9.1 | 9.0 | [5.0; 13.0] |
| Ismaila et al. (2016) | 35 | 22.5 | 8.5 | 22.5 | [19.7; 25.3] |
| **Random Effect model** | **249** | | | **10.5** | **[6.9; 14.1]** |

**Table S4:** BC ASSR thresholds in NH infants at 500 and 1000 Hz.

| Study | Total | Mean | SD | MRAW | 95%-CI |
|---|---|---|---|---|---|
| **Freq = 2000 Hz** | | | | | |
| Small and Stapells (2006) | 29 | 37.3 | 15.9 | 37.3 | [31.5; 43.1] |
| Small and Stapells (2006) | 14 | 26.4 | 6.3 | 26.4 | [23.1; 29.7] |
| Small et al. (2007) | 10 | 26.0 | 9.7 | 26.0 | [20.0; 32.0] |
| Small et al. (2007) | 13 | 34.6 | 15.1 | 34.6 | [26.4; 42.8] |
| Small et al. (2007) | 13 | 30.0 | 5.8 | 30.0 | [26.8; 33.2] |
| Small and Stapells (2008a) | 35 | 26.6 | 10.2 | 26.6 | [23.2; 30.0] |
| Small and Stapells (2008a) | 13 | 26.6 | 8.6 | 26.6 | [21.9; 31.2] |
| Small and Stapells (2008b) | 12 | 20.0 | 12.8 | 20.0 | [12.8; 27.2] |
| Swanepoel et al. (2008) | 21 | 23.6 | 6.5 | 23.6 | [20.8; 26.4] |
| Small and Hu (2011) | 22 | 28.6 | 8.9 | 28.6 | [24.8; 32.3] |
| Small and Hu (2011) | 10 | 27.0 | 13.6 | 27.0 | [18.6; 35.4] |
| Casey and Small (2014) | 20 | 20.5 | 10.5 | 20.5 | [15.9; 25.1] |
| Ismaila et al. (2016) | 35 | 20.0 | 6.5 | 20.0 | [17.8; 22.2] |
| Valeriote and Small (2024) | 21 | 21.0 | 14.0 | 21.0 | [15.0; 27.0] |
| **Random Effect model** | **268** | | | **26.1** | **[23.5; 28.6]** |
| **Freq = 4000 Hz** | | | | | |
| Small and Stapells (2006) | 29 | 32.5 | 12.7 | 32.5 | [27.9; 37.1] |
| Small and Stapells (2006) | 14 | 22.1 | 8.0 | 22.1 | [17.9; 26.3] |
| Small et al. (2007) | 10 | 13.0 | 11.6 | 13.0 | [5.8; 20.2] |
| Small et al. (2007) | 15 | 33.3 | 15.0 | 33.3 | [25.7; 40.9] |
| Small et al. (2007) | 13 | 16.2 | 9.6 | 16.2 | [11.0; 21.4] |
| Small and Stapells (2008a) | 35 | 13.9 | 10.5 | 13.9 | [10.4; 17.3] |
| Small and Stapells (2008a) | 13 | 13.4 | 9.5 | 13.4 | [8.2; 18.5] |
| Small and Stapells (2008b) | 12 | 9.2 | 7.9 | 9.2 | [4.7; 13.7] |
| Swanepoel et al. (2008) | 21 | 25.5 | 7.6 | 25.5 | [22.2; 28.8] |
| Small and Hu (2011) | 22 | 21.1 | 11.0 | 21.1 | [16.5; 25.7] |
| Small and Hu (2011) | 10 | 19.6 | 10.8 | 19.6 | [12.9; 26.3] |
| Casey and Small (2014) | 20 | 14.5 | 11.4 | 14.5 | [9.5; 19.5] |
| Ismaila et al. (2016) | 35 | 25.0 | 6.1 | 25.0 | [23.0; 27.0] |
| **Random Effect model** | **249** | | | **19.9** | **[15.9; 23.9]** |

**Table S5:** BC ASSR thresholds in NH infants at 2000 and 4000 Hz.

| Study | Total | Mean | SD | MRAW | 95%-CI |
|---|---|---|---|---|---|
| Swanepoel et al. (2008) | 35.0 | 19.4 | 8.5 | 19.4 | [16.6; 22.2] |
| Ismaila et al. (2016) | 35.0 | 24.0 | 6.4 | 24.0 | [21.9; 26.1] |
| Valeriote and Small (2024) | 15.0 | 15.3 | 12.6 | 15.3 | [8.9; 21.7] |
| **Random Effect model** | **85** | | | **20.3** | **[15.6; 24.9]** |

**Table S6:** BC ASSR thresholds in CHL infants at 500 Hz.

| Paper | N | AgeMean | AgeMetric | AgeGroup | HearingGroup | Test | Metric | Freq | Thr |
|---|---|---|---|---|---|---|---|---|---|
| Lins et al (1996) | 6.00 | | year | Adults | NH | B | M | 500 | 16.00 |
| Lins et al (1996) | 6.00 | | year | Adults | NH | B | M | 1000 | 13.00 |
| Lins et al (1996) | 6.00 | | year | Adults | NH | B | M | 2000 | 24.00 |
| Lins et al (1996) | 6.00 | | year | Adults | NH | B | M | 4000 | 16.00 |
| Lins et al (1996) | 6.00 | | year | Adults | NH | B | S | 500 | 8.00 |
| Lins et al (1996) | 6.00 | | year | Adults | NH | B | S | 1000 | 5.00 |
| Lins et al (1996) | 6.00 | | year | Adults | NH | B | S | 2000 | 4.00 |
| Lins et al (1996) | 6.00 | | year | Adults | NH | B | S | 4000 | 4.00 |
| Lins et al (1996) | 6.00 | | year | Adults | NH | A | M | 500 | 26.00 |
| Lins et al (1996) | 6.00 | | year | Adults | NH | A | M | 1000 | 28.00 |
| Lins et al (1996) | 6.00 | | year | Adults | NH | A | M | 2000 | 33.00 |
| Lins et al (1996) | 6.00 | | year | Adults | NH | A | M | 4000 | 26.00 |
| Lins et al (1996) | 6.00 | | year | Adults | NH | A | S | 500 | 6.00 |
| Lins et al (1996) | 6.00 | | year | Adults | NH | A | S | 1000 | 10.00 |
| Lins et al (1996) | 6.00 | | year | Adults | NH | A | S | 2000 | 7.00 |
| Lins et al (1996) | 6.00 | | year | Adults | NH | A | S | 4000 | 11.00 |
| Small & Stapells (2005) | 10.00 | | year | Adults | NH | B | M | 500 | 4.90 |
| Small & Stapells (2005) | 10.00 | | year | Adults | NH | B | S | 500 | 7.70 |
| Small & Stapells (2005) | 10.00 | | year | Adults | NH | B | M | 1000 | -3.10 |
| Small & Stapells (2005) | 10.00 | | year | Adults | NH | B | S | 1000 | 4.90 |
| Small & Stapells (2005) | 10.00 | | year | Adults | NH | B | M | 2000 | 4.40 |
| Small & Stapells (2005) | 10.00 | | year | Adults | NH | B | S | 2000 | 6.30 |
| Small & Stapells (2005) | 10.00 | | year | Adults | NH | B | M | 4000 | -3.60 |
| Small & Stapells (2005) | 10.00 | | year | Adults | NH | B | S | 4000 | 6.30 |
| Small & Stapells (2005) | 10.00 | | year | Adults | NH | A | M | 500 | 22.00 |
| Small & Stapells (2005) | 10.00 | | year | Adults | NH | A | S | 500 | 11.40 |
| Small & Stapells (2005) | 10.00 | | year | Adults | NH | A | M | 1000 | 26.00 |
| Small & Stapells (2005) | 10.00 | | year | Adults | NH | A | S | 1000 | 13.50 |
| Small & Stapells (2005) | 10.00 | | year | Adults | NH | A | M | 2000 | 18.00 |
| Small & Stapells (2005) | 10.00 | | year | Adults | NH | A | S | 2000 | 7.90 |
| Small & Stapells (2005) | 10.00 | | year | Adults | NH | A | M | 4000 | 18.00 |
| Small & Stapells (2005) | 10.00 | | year | Adults | NH | A | S | 4000 | 11.40 |
| Small & Stapells (2006) | 29.00 | 34.50 | wk pca | Infants | NH | A | M | 500 | 16.20 |
| Small & Stapells (2006) | 29.00 | 34.50 | wk pca | Infants | NH | A | S | 500 | 10.80 |
| Small & Stapells (2006) | 29.00 | 34.50 | wk pca | Infants | NH | A | M | 1000 | 15.50 |
| Small & Stapells (2006) | 29.00 | 34.50 | wk pca | Infants | NH | A | S | 1000 | 10.20 |
| Small & Stapells (2006) | 29.00 | 34.50 | wk pca | Infants | NH | A | M | 2000 | 37.30 |
| Small & Stapells (2006) | 29.00 | 34.50 | wk pca | Infants | NH | A | S | 2000 | 15.90 |
| Small & Stapells (2006) | 29.00 | 34.50 | wk pca | Infants | NH | A | M | 4000 | 32.50 |
| Small & Stapells (2006) | 29.00 | 34.50 | wk pca | Infants | NH | A | S | 4000 | 12.70 |
| Small & Stapells (2006) | 14.00 | 17.00 | week | Infants | NH | A | M | 500 | 13.60 |
| Small & Stapells (2006) | 14.00 | 17.00 | week | Infants | NH | A | S | 500 | 13.40 |
| Small & Stapells (2006) | 14.00 | 17.00 | week | Infants | NH | A | M | 1000 | 2.10 |
| Small & Stapells (2006) | 14.00 | 17.00 | week | Infants | NH | A | S | 1000 | 7.00 |
| Small & Stapells (2006) | 14.00 | 17.00 | week | Infants | NH | A | M | 2000 | 26.40 |
| Small & Stapells (2006) | 14.00 | 17.00 | week | Infants | NH | A | S | 2000 | 6.30 |
| Small & Stapells (2006) | 14.00 | 17.00 | week | Infants | NH | A | M | 4000 | 22.10 |
| Small & Stapells (2006) | 14.00 | 17.00 | week | Infants | NH | A | S | 4000 | 8.00 |
| Small et al (2007) | 10.00 | 17.00 | week | Infants | NH | A | M | 500 | 14.00 |
| Small et al (2007) | 10.00 | 17.00 | week | Infants | NH | A | S | 500 | 14.30 |
| Small et al (2007) | 10.00 | 17.00 | week | Infants | NH | A | M | 1000 | 6.00 |
| Small et al (2007) | 10.00 | 17.00 | week | Infants | NH | A | S | 1000 | 8.43 |
| Small et al (2007) | 10.00 | 17.00 | week | Infants | NH | A | M | 2000 | 26.00 |
| Small et al (2007) | 10.00 | 17.00 | week | Infants | NH | A | S | 2000 | 9.70 |
| Small et al (2007) | 10.00 | 17.00 | week | Infants | NH | A | M | 4000 | 13.00 |
| Small et al (2007) | 10.00 | 17.00 | week | Infants | NH | A | S | 4000 | 11.60 |
| Small et al (2007) | 15.00 | 34.50 | wk pca | Infants | NH | A | M | 500 | 16.00 |
| Small et al (2007) | 15.00 | 34.50 | wk pca | Infants | NH | A | S | 500 | 11.80 |
| Small et al (2007) | 15.00 | 34.50 | wk pca | Infants | NH | A | M | 1000 | 16.70 |
| Small et al (2007) | 15.00 | 34.50 | wk pca | Infants | NH | A | S | 1000 | 9.00 |
| Small et al (2007) | 13.00 | 34.50 | wk pca | Infants | NH | A | M | 2000 | 34.60 |
| Small et al (2007) | 13.00 | 34.50 | wk pca | Infants | NH | A | S | 2000 | 15.10 |
| Small et al (2007) | 15.00 | 34.50 | wk pca | Infants | NH | A | M | 4000 | 33.30 |
| Small et al (2007) | 15.00 | 34.50 | wk pca | Infants | NH | A | S | 4000 | 15.00 |
| Small et al (2007) | 13.00 | 15.00 | week | Infants | NH | A | M | 500 | 18.50 |
| Small et al (2007) | 13.00 | 15.00 | week | Infants | NH | A | S | 500 | 14.10 |
| Small et al (2007) | 13.00 | 15.00 | week | Infants | NH | A | M | 1000 | 3.10 |
| Small et al (2007) | 13.00 | 15.00 | week | Infants | NH | A | S | 1000 | 8.60 |
| Small et al (2007) | 13.00 | 15.00 | week | Infants | NH | A | M | 2000 | 30.00 |

| | | | | | | | | | |
|---|---|---|---|---|---|---|---|---|---|
| Small et al (2007) | 13.00 | 15.00 | week | Infants | NH | A | S | 2000 | 5.80 |
| Small et al (2007) | 13.00 | 15.00 | week | Infants | NH | A | M | 4000 | 16.20 |
| Small et al (2007) | 13.00 | 15.00 | week | Infants | NH | A | S | 4000 | 9.60 |
| Small & Stapells (2008a) | 35.00 | 16.00 | week | Infants | NH | A | M | 500 | 14.22 |
| Small & Stapells (2008a) | 35.00 | 16.00 | week | Infants | NH | A | S | 500 | 12.87 |
| Small & Stapells (2008a) | 35.00 | 16.00 | week | Infants | NH | A | M | 1000 | 5.25 |
| Small & Stapells (2008a) | 35.00 | 16.00 | week | Infants | NH | A | S | 1000 | 7.69 |
| Small & Stapells (2008a) | 35.00 | 16.00 | week | Infants | NH | A | M | 2000 | 26.61 |
| Small & Stapells (2008a) | 35.00 | 16.00 | week | Infants | NH | A | S | 2000 | 10.15 |
| Small & Stapells (2008a) | 35.00 | 16.00 | week | Infants | NH | A | M | 4000 | 13.86 |
| Small & Stapells (2008a) | 35.00 | 16.00 | week | Infants | NH | A | S | 4000 | 10.52 |
| Small & Stapells (2008a) | 13.00 | 18.20 | months | Infants | NH | A | M | 500 | 22.55 |
| Small & Stapells (2008a) | 13.00 | 18.20 | months | Infants | NH | A | S | 500 | 10.84 |
| Small & Stapells (2008a) | 13.00 | 18.20 | months | Infants | NH | A | M | 1000 | 13.32 |
| Small & Stapells (2008a) | 13.00 | 18.20 | months | Infants | NH | A | S | 1000 | 6.28 |
| Small & Stapells (2008a) | 13.00 | 18.20 | months | Infants | NH | A | M | 2000 | 26.57 |
| Small & Stapells (2008a) | 13.00 | 18.20 | months | Infants | NH | A | S | 2000 | 8.60 |
| Small & Stapells (2008a) | 13.00 | 18.20 | months | Infants | NH | A | M | 4000 | 13.38 |
| Small & Stapells (2008a) | 13.00 | 18.20 | months | Infants | NH | A | S | 4000 | 9.51 |
| Small & Stapells (2008a) | 18.00 | 22.90 | year | Adults | NH | A | M | 500 | 30.94 |
| Small & Stapells (2008a) | 18.00 | 22.90 | year | Adults | NH | A | S | 500 | 15.12 |
| Small & Stapells (2008a) | 18.00 | 22.90 | year | Adults | NH | A | M | 1000 | 24.32 |
| Small & Stapells (2008a) | 18.00 | 22.90 | year | Adults | NH | A | S | 1000 | 13.78 |
| Small & Stapells (2008a) | 18.00 | 22.90 | year | Adults | NH | A | M | 2000 | 20.54 |
| Small & Stapells (2008a) | 18.00 | 22.90 | year | Adults | NH | A | S | 2000 | 7.72 |
| Small & Stapells (2008a) | 18.00 | 22.90 | year | Adults | NH | A | M | 4000 | 16.27 |
| Small & Stapells (2008a) | 18.00 | 22.90 | year | Adults | NH | A | S | 4000 | 10.90 |
| Small & Stapells (2008b) | 12.00 | 21.00 | week | Infants | NH | A | M | 500 | 12.50 |
| Small & Stapells (2008b) | 12.00 | 21.00 | week | Infants | NH | A | S | 500 | 12.90 |
| Small & Stapells (2008b) | 12.00 | 21.00 | week | Infants | NH | A | M | 1000 | 5.00 |
| Small & Stapells (2008b) | 12.00 | 21.00 | week | Infants | NH | A | S | 1000 | 5.20 |
| Small & Stapells (2008b) | 12.00 | 21.00 | week | Infants | NH | A | M | 2000 | 20.00 |
| Small & Stapells (2008b) | 12.00 | 21.00 | week | Infants | NH | A | S | 2000 | 12.80 |
| Small & Stapells (2008b) | 12.00 | 21.00 | week | Infants | NH | A | M | 4000 | 9.20 |
| Small & Stapells (2008b) | 12.00 | 21.00 | week | Infants | NH | A | S | 4000 | 7.90 |
| Small & Stapells (2008b) | 8.00 | 23.00 | year | Adults | NH | A | M | 500 | 31.30 |
| Small & Stapells (2008b) | 8.00 | 23.00 | year | Adults | NH | A | S | 500 | 6.40 |
| Small & Stapells (2008b) | 8.00 | 23.00 | year | Adults | NH | A | M | 1000 | 17.50 |
| Small & Stapells (2008b) | 8.00 | 23.00 | year | Adults | NH | A | S | 1000 | 12.80 |
| Small & Stapells (2008b) | 8.00 | 23.00 | year | Adults | NH | A | M | 2000 | 20.00 |
| Small & Stapells (2008b) | 8.00 | 23.00 | year | Adults | NH | A | S | 2000 | 7.60 |
| Small & Stapells (2008b) | 8.00 | 23.00 | year | Adults | NH | A | M | 4000 | 10.00 |
| Small & Stapells (2008b) | 8.00 | 23.00 | year | Adults | NH | A | S | 4000 | 10.70 |
| Swanepoel et al (2008) | 23.00 | 2.20 | year | Infants | SNHL | A | M | 500 | 53.00 |
| Swanepoel et al (2008) | 23.00 | 2.20 | year | Infants | SNHL | A | S | 500 | 4.90 |
| Swanepoel et al (2008) | 23.00 | 2.20 | year | Infants | SNHL | A | M | 1000 | 61.10 |
| Swanepoel et al (2008) | 23.00 | 2.20 | year | Infants | SNHL | A | S | 1000 | 2.10 |
| Swanepoel et al (2008) | 23.00 | 2.20 | year | Infants | SNHL | A | M | 2000 | 68.00 |
| Swanepoel et al (2008) | 23.00 | 2.20 | year | Infants | SNHL | A | S | 2000 | 3.60 |
| Swanepoel et al (2008) | 22.00 | 2.20 | year | Infants | SNHL | A | M | 4000 | 68.40 |
| Swanepoel et al (2008) | 22.00 | 2.20 | year | Infants | SNHL | A | S | 4000 | 2.80 |
| Swanepoel et al (2008) | 21.00 | 3.60 | year | Infants | NH | A | M | 500 | 17.90 |
| Swanepoel et al (2008) | 21.00 | 3.60 | year | Infants | NH | A | S | 500 | 6.80 |
| Swanepoel et al (2008) | 21.00 | 3.60 | year | Infants | NH | A | M | 1000 | 16.00 |
| Swanepoel et al (2008) | 21.00 | 3.60 | year | Infants | NH | A | S | 1000 | 11.40 |
| Swanepoel et al (2008) | 21.00 | 3.60 | year | Infants | NH | A | M | 2000 | 23.60 |
| Swanepoel et al (2008) | 21.00 | 3.60 | year | Infants | NH | A | S | 2000 | 6.50 |
| Swanepoel et al (2008) | 21.00 | 3.60 | year | Infants | NH | A | M | 4000 | 25.50 |
| Swanepoel et al (2008) | 21.00 | 3.60 | year | Infants | NH | A | S | 4000 | 7.60 |
| Swanepoel et al (2008) | 13.00 | 2.50 | year | Infants | SNHL | A | M | 500 | 36.50 |
| Swanepoel et al (2008) | 13.00 | 2.50 | year | Infants | SNHL | A | S | 500 | 6.60 |
| Swanepoel et al (2008) | 13.00 | 2.50 | year | Infants | SNHL | A | M | 1000 | 41.50 |
| Swanepoel et al (2008) | 13.00 | 2.50 | year | Infants | SNHL | A | S | 1000 | 9.90 |
| Swanepoel et al (2008) | 13.00 | 2.50 | year | Infants | SNHL | A | M | 2000 | 56.50 |
| Swanepoel et al (2008) | 13.00 | 2.50 | year | Infants | SNHL | A | S | 2000 | 10.90 |
| Swanepoel et al (2008) | 13.00 | 2.50 | year | Infants | SNHL | A | M | 4000 | 55.40 |
| Swanepoel et al (2008) | 13.00 | 2.50 | year | Infants | SNHL | A | S | 4000 | 13.00 |
| Swanepoel et al (2008) | 35.00 | 2.70 | year | Infants | CHL | A | M | 500 | 19.40 |
| Swanepoel et al (2008) | 35.00 | 2.70 | year | Infants | CHL | A | S | 500 | 8.50 |
| Swanepoel et al (2008) | 35.00 | 2.70 | year | Infants | CHL | A | M | 1000 | 25.00 |
| Swanepoel et al (2008) | 35.00 | 2.70 | year | Infants | CHL | A | S | 1000 | 11.30 |
| Swanepoel et al (2008) | 35.00 | 2.70 | year | Infants | CHL | A | M | 2000 | 24.30 |

| | | | | | | | | | |
|---|---|---|---|---|---|---|---|---|---|
| Swanepoel et al (2008) | 35.00 | 2.70 | year | Infants | CHL | A | S | 2000 | 8.50 |
| Swanepoel et al (2008) | 35.00 | 2.70 | year | Infants | CHL | A | M | 4000 | 25.30 |
| Swanepoel et al (2008) | 35.00 | 2.70 | year | Infants | CHL | A | S | 4000 | 11.90 |
| Ishida et al (2011) | 14.00 | 26.80 | year | Adults | NH | B | M | 500 | 1.10 |
| Ishida et al (2011) | 14.00 | 26.80 | year | Adults | NH | B | S | 500 | 8.40 |
| Ishida et al (2011) | 15.00 | 26.80 | year | Adults | NH | B | M | 1000 | 1.70 |
| Ishida et al (2011) | 15.00 | 26.80 | year | Adults | NH | B | S | 1000 | 5.90 |
| Ishida et al (2011) | 15.00 | 26.80 | year | Adults | NH | B | M | 2000 | 9.30 |
| Ishida et al (2011) | 15.00 | 26.80 | year | Adults | NH | B | S | 2000 | 6.50 |
| Ishida et al (2011) | 16.00 | 26.80 | year | Adults | NH | B | M | 4000 | 14.40 |
| Ishida et al (2011) | 16.00 | 26.80 | year | Adults | NH | B | S | 4000 | 6.00 |
| Ishida et al (2011) | 14.00 | 26.80 | year | Adults | NH | A | M | 500 | 20.70 |
| Ishida et al (2011) | 14.00 | 26.80 | year | Adults | NH | A | S | 500 | 10.70 |
| Ishida et al (2011) | 15.00 | 26.80 | year | Adults | NH | A | M | 1000 | 14.00 |
| Ishida et al (2011) | 15.00 | 26.80 | year | Adults | NH | A | S | 1000 | 6.30 |
| Ishida et al (2011) | 15.00 | 26.80 | year | Adults | NH | A | M | 2000 | 22.00 |
| Ishida et al (2011) | 15.00 | 26.80 | year | Adults | NH | A | S | 2000 | 7.80 |
| Ishida et al (2011) | 16.00 | 26.80 | year | Adults | NH | A | M | 4000 | 27.50 |
| Ishida et al (2011) | 16.00 | 26.80 | year | Adults | NH | A | S | 4000 | 8.60 |
| Ishida et al (2011) | 10.00 | 24.60 | year | Adults | NH | B | M | 500 | 8.00 |
| Ishida et al (2011) | 10.00 | 24.60 | year | Adults | NH | B | S | 500 | 5.40 |
| Ishida et al (2011) | 10.00 | 24.60 | year | Adults | NH | B | M | 1000 | 4.00 |
| Ishida et al (2011) | 10.00 | 24.60 | year | Adults | NH | B | S | 1000 | 3.90 |
| Ishida et al (2011) | 10.00 | 24.60 | year | Adults | NH | B | M | 2000 | 5.00 |
| Ishida et al (2011) | 10.00 | 24.60 | year | Adults | NH | B | S | 2000 | 5.30 |
| Ishida et al (2011) | 10.00 | 24.60 | year | Adults | NH | B | M | 4000 | 3.00 |
| Ishida et al (2011) | 10.00 | 24.60 | year | Adults | NH | B | S | 4000 | 6.30 |
| Ishida et al (2011) | 10.00 | 24.60 | year | Adults | NH | A | M | 500 | 21.70 |
| Ishida et al (2011) | 10.00 | 24.60 | year | Adults | NH | A | S | 500 | 9.40 |
| Ishida et al (2011) | 10.00 | 24.60 | year | Adults | NH | A | M | 1000 | 15.60 |
| Ishida et al (2011) | 10.00 | 24.60 | year | Adults | NH | A | S | 1000 | 12.90 |
| Ishida et al (2011) | 10.00 | 24.60 | year | Adults | NH | A | M | 2000 | 15.90 |
| Ishida et al (2011) | 10.00 | 24.60 | year | Adults | NH | A | S | 2000 | 9.50 |
| Ishida et al (2011) | 10.00 | 24.60 | year | Adults | NH | A | M | 4000 | 11.00 |
| Ishida et al (2011) | 10.00 | 24.60 | year | Adults | NH | A | S | 4000 | 3.20 |
| Ishida et al (2011) | 39.00 | 46.00 | year | Adults | SNHL | B | M | 500 | 31.80 |
| Ishida et al (2011) | 39.00 | 46.00 | year | Adults | SNHL | B | S | 500 | 15.70 |
| Ishida et al (2011) | 39.00 | 46.00 | year | Adults | SNHL | B | M | 1000 | 34.10 |
| Ishida et al (2011) | 39.00 | 46.00 | year | Adults | SNHL | B | S | 1000 | 17.40 |
| Ishida et al (2011) | 39.00 | 46.00 | year | Adults | SNHL | B | M | 2000 | 43.50 |
| Ishida et al (2011) | 39.00 | 46.00 | year | Adults | SNHL | B | S | 2000 | 14.70 |
| Ishida et al (2011) | 39.00 | 46.00 | year | Adults | SNHL | B | M | 4000 | 48.20 |
| Ishida et al (2011) | 39.00 | 46.00 | year | Adults | SNHL | B | S | 4000 | 13.70 |
| Ishida et al (2011) | 39.00 | 46.00 | year | Adults | SNHL | A | M | 500 | 48.70 |
| Ishida et al (2011) | 39.00 | 46.00 | year | Adults | SNHL | A | S | 500 | 12.80 |
| Ishida et al (2011) | 39.00 | 46.00 | year | Adults | SNHL | A | M | 1000 | 44.60 |
| Ishida et al (2011) | 39.00 | 46.00 | year | Adults | SNHL | A | S | 1000 | 14.10 |
| Ishida et al (2011) | 39.00 | 46.00 | year | Adults | SNHL | A | M | 2000 | 52.60 |
| Ishida et al (2011) | 39.00 | 46.00 | year | Adults | SNHL | A | S | 2000 | 12.30 |
| Ishida et al (2011) | 39.00 | 46.00 | year | Adults | SNHL | A | M | 4000 | 53.10 |
| Ishida et al (2011) | 39.00 | 46.00 | year | Adults | SNHL | A | S | 4000 | 11.70 |
| Small & Hu (2011) | 22.00 | 2.10 | months | Infants | NH | A | M | 500 | 18.32 |
| Small & Hu (2011) | 22.00 | 2.10 | months | Infants | NH | A | S | 500 | 11.78 |
| Small & Hu (2011) | 22.00 | 2.10 | months | Infants | NH | A | M | 1000 | 5.71 |
| Small & Hu (2011) | 22.00 | 2.10 | months | Infants | NH | A | S | 1000 | 8.60 |
| Small & Hu (2011) | 22.00 | 2.10 | months | Infants | NH | A | M | 2000 | 28.55 |
| Small & Hu (2011) | 22.00 | 2.10 | months | Infants | NH | A | S | 2000 | 8.90 |
| Small & Hu (2011) | 22.00 | 2.10 | months | Infants | NH | A | M | 4000 | 21.14 |
| Small & Hu (2011) | 22.00 | 2.10 | months | Infants | NH | A | S | 4000 | 11.00 |
| Small & Hu (2011) | 10.00 | 15.30 | months | Infants | NH | A | M | 500 | 22.32 |
| Small & Hu (2011) | 10.00 | 15.30 | months | Infants | NH | A | S | 500 | 10.84 |
| Small & Hu (2011) | 10.00 | 15.30 | months | Infants | NH | A | M | 1000 | 16.90 |
| Small & Hu (2011) | 10.00 | 15.30 | months | Infants | NH | A | S | 1000 | 8.25 |
| Small & Hu (2011) | 10.00 | 15.30 | months | Infants | NH | A | M | 2000 | 26.99 |
| Small & Hu (2011) | 10.00 | 15.30 | months | Infants | NH | A | S | 2000 | 13.58 |
| Small & Hu (2011) | 10.00 | 15.30 | months | Infants | NH | A | M | 4000 | 19.58 |
| Small & Hu (2011) | 10.00 | 15.30 | months | Infants | NH | A | S | 4000 | 10.77 |
| Small & Hu (2011) | 20.00 | 26.50 | year | Adults | NH | B | M | 500 | -0.70 |
| Small & Hu (2011) | 20.00 | 26.50 | year | Adults | NH | B | S | 500 | 6.60 |
| Small & Hu (2011) | 20.00 | 26.50 | year | Adults | NH | B | M | 1000 | 5.12 |
| Small & Hu (2011) | 20.00 | 26.50 | year | Adults | NH | B | S | 1000 | 5.65 |
| Small & Hu (2011) | 20.00 | 26.50 | year | Adults | NH | B | M | 2000 | 0.69 |

| | | | | | | | | | |
|---|---|---|---|---|---|---|---|---|---|
| Small & Hu (2011) | 20.00 | 26.50 | year | Adults | NH | B | S | 2000 | 5.86 |
| Small & Hu (2011) | 20.00 | 26.50 | year | Adults | NH | B | M | 4000 | 3.58 |
| Small & Hu (2011) | 20.00 | 26.50 | year | Adults | NH | B | S | 4000 | 6.08 |
| Small & Hu (2011) | 20.00 | 26.50 | year | Adults | NH | A | M | 500 | 23.27 |
| Small & Hu (2011) | 20.00 | 26.50 | year | Adults | NH | A | S | 500 | 14.14 |
| Small & Hu (2011) | 20.00 | 26.50 | year | Adults | NH | A | M | 1000 | 16.90 |
| Small & Hu (2011) | 20.00 | 26.50 | year | Adults | NH | A | S | 1000 | 12.02 |
| Small & Hu (2011) | 20.00 | 26.50 | year | Adults | NH | A | M | 2000 | 18.41 |
| Small & Hu (2011) | 20.00 | 26.50 | year | Adults | NH | A | S | 2000 | 6.55 |
| Small & Hu (2011) | 20.00 | 26.50 | year | Adults | NH | A | M | 4000 | 14.04 |
| Small & Hu (2011) | 20.00 | 26.50 | year | Adults | NH | A | S | 4000 | 9.83 |
| Casey & Small (2014) | 19.00 | 10.50 | months | Infants | NH | B | M | 500 | -0.50 |
| Casey & Small (2014) | 19.00 | 10.50 | months | Infants | NH | B | S | 500 | 9.10 |
| Casey & Small (2014) | 4.00 | 10.50 | months | Infants | NH | B | M | 1000 | 2.50 |
| Casey & Small (2014) | 4.00 | 10.50 | months | Infants | NH | B | S | 1000 | 12.60 |
| Casey & Small (2014) | 20.00 | 10.50 | months | Infants | NH | B | M | 2000 | 2.50 |
| Casey & Small (2014) | 20.00 | 10.50 | months | Infants | NH | B | S | 2000 | 8.50 |
| Casey & Small (2014) | 18.00 | 10.50 | months | Infants | NH | B | M | 4000 | 5.60 |
| Casey & Small (2014) | 18.00 | 10.50 | months | Infants | NH | B | S | 4000 | 7.00 |
| Casey & Small (2014) | 19.00 | 10.50 | months | Infants | NH | A | M | 500 | 11.10 |
| Casey & Small (2014) | 19.00 | 10.50 | months | Infants | NH | A | S | 500 | 11.00 |
| Casey & Small (2014) | 20.00 | 10.50 | months | Infants | NH | A | M | 1000 | 9.00 |
| Casey & Small (2014) | 20.00 | 10.50 | months | Infants | NH | A | S | 1000 | 9.10 |
| Casey & Small (2014) | 20.00 | 10.50 | months | Infants | NH | A | M | 2000 | 20.50 |
| Casey & Small (2014) | 20.00 | 10.50 | months | Infants | NH | A | S | 2000 | 10.50 |
| Casey & Small (2014) | 20.00 | 10.50 | months | Infants | NH | A | M | 4000 | 14.50 |
| Casey & Small (2014) | 20.00 | 10.50 | months | Infants | NH | A | S | 4000 | 11.40 |
| Casey & Small (2014) | 8.00 | 29.90 | year | Adults | NH | B | M | 500 | 1.30 |
| Casey & Small (2014) | 8.00 | 29.90 | year | Adults | NH | B | S | 500 | 6.40 |
| Casey & Small (2014) | 8.00 | 29.90 | year | Adults | NH | B | M | 1000 | 1.30 |
| Casey & Small (2014) | 8.00 | 29.90 | year | Adults | NH | B | S | 1000 | 4.40 |
| Casey & Small (2014) | 8.00 | 29.90 | year | Adults | NH | B | M | 2000 | 2.50 |
| Casey & Small (2014) | 8.00 | 29.90 | year | Adults | NH | B | S | 2000 | 4.60 |
| Casey & Small (2014) | 8.00 | 29.90 | year | Adults | NH | B | M | 4000 | 0.00 |
| Casey & Small (2014) | 8.00 | 29.90 | year | Adults | NH | B | S | 4000 | 6.00 |
| Casey & Small (2014) | 11.00 | 29.90 | year | Adults | NH | A | M | 500 | 20.00 |
| Casey & Small (2014) | 11.00 | 29.90 | year | Adults | NH | A | S | 500 | 8.90 |
| Casey & Small (2014) | 11.00 | 29.90 | year | Adults | NH | A | M | 1000 | 15.50 |
| Casey & Small (2014) | 11.00 | 29.90 | year | Adults | NH | A | S | 1000 | 12.10 |
| Casey & Small (2014) | 11.00 | 29.90 | year | Adults | NH | A | M | 2000 | 12.70 |
| Casey & Small (2014) | 11.00 | 29.90 | year | Adults | NH | A | S | 2000 | 7.90 |
| Casey & Small (2014) | 11.00 | 29.90 | year | Adults | NH | A | M | 4000 | 8.50 |
| Casey & Small (2014) | 11.00 | 29.90 | year | Adults | NH | A | S | 4000 | 10.40 |
| Ismaila et al (2016) | 35.00 | 4.50 | year | Infants | NH | B | M | 500 | 13.00 |
| Ismaila et al (2016) | 35.00 | 4.50 | year | Infants | NH | B | S | 500 | 2.50 |
| Ismaila et al (2016) | 35.00 | 4.50 | year | Infants | NH | B | M | 1000 | 15.00 |
| Ismaila et al (2016) | 35.00 | 4.50 | year | Infants | NH | B | S | 1000 | 3.20 |
| Ismaila et al (2016) | 35.00 | 4.50 | year | Infants | NH | B | M | 2000 | 14.50 |
| Ismaila et al (2016) | 35.00 | 4.50 | year | Infants | NH | B | S | 2000 | 2.24 |
| Ismaila et al (2016) | 35.00 | 4.50 | year | Infants | NH | B | M | 4000 | 14.50 |
| Ismaila et al (2016) | 35.00 | 4.50 | year | Infants | NH | B | S | 4000 | 3.20 |
| Ismaila et al (2016) | 35.00 | 4.50 | year | Infants | NH | A | M | 500 | 23.50 |
| Ismaila et al (2016) | 35.00 | 4.50 | year | Infants | NH | A | S | 500 | 12.26 |
| Ismaila et al (2016) | 35.00 | 4.50 | year | Infants | NH | A | M | 1000 | 22.50 |
| Ismaila et al (2016) | 35.00 | 4.50 | year | Infants | NH | A | S | 1000 | 8.51 |
| Ismaila et al (2016) | 35.00 | 4.50 | year | Infants | NH | A | M | 2000 | 20.00 |
| Ismaila et al (2016) | 35.00 | 4.50 | year | Infants | NH | A | S | 2000 | 6.49 |
| Ismaila et al (2016) | 35.00 | 4.50 | year | Infants | NH | A | M | 4000 | 25.00 |
| Ismaila et al (2016) | 35.00 | 4.50 | year | Infants | NH | A | S | 4000 | 6.07 |
| Ismaila et al (2016) | 36.00 | 4.50 | year | Infants | SNHL | B | M | 500 | 36.25 |
| Ismaila et al (2016) | 36.00 | 4.50 | year | Infants | SNHL | B | S | 500 | 4.50 |
| Ismaila et al (2016) | 36.00 | 4.50 | year | Infants | SNHL | B | M | 1000 | 35.75 |
| Ismaila et al (2016) | 36.00 | 4.50 | year | Infants | SNHL | B | S | 1000 | 4.67 |
| Ismaila et al (2016) | 36.00 | 4.50 | year | Infants | SNHL | B | M | 2000 | 42.75 |
| Ismaila et al (2016) | 36.00 | 4.50 | year | Infants | SNHL | B | S | 2000 | 4.99 |
| Ismaila et al (2016) | 36.00 | 4.50 | year | Infants | SNHL | B | M | 4000 | 46.75 |
| Ismaila et al (2016) | 36.00 | 4.50 | year | Infants | SNHL | B | S | 4000 | 2.94 |
| Ismaila et al (2016) | 36.00 | 4.50 | year | Infants | SNHL | A | M | 500 | 52.50 |
| Ismaila et al (2016) | 36.00 | 4.50 | year | Infants | SNHL | A | S | 500 | 6.39 |
| Ismaila et al (2016) | 36.00 | 4.50 | year | Infants | SNHL | A | M | 1000 | 41.50 |
| Ismaila et al (2016) | 36.00 | 4.50 | year | Infants | SNHL | A | S | 1000 | 6.71 |
| Ismaila et al (2016) | 36.00 | 4.50 | year | Infants | SNHL | A | M | 2000 | 55.00 |

| | | | | | | | | | |
|---|---|---|---|---|---|---|---|---|---|
| Ismaila et al (2016) | 36.00 | 4.50 | year | Infants | SNHL | A | S | 2000 | 5.13 |
| Ismaila et al (2016) | 36.00 | 4.50 | year | Infants | SNHL | A | M | 4000 | 58.50 |
| Ismaila et al (2016) | 36.00 | 4.50 | year | Infants | SNHL | A | S | 4000 | 3.66 |
| Ismaila et al (2016) | 35.00 | 4.50 | year | Infants | CHL | B | M | 500 | 11.50 |
| Ismaila et al (2016) | 35.00 | 4.50 | year | Infants | CHL | B | S | 500 | 2.30 |
| Ismaila et al (2016) | 35.00 | 4.50 | year | Infants | CHL | B | M | 1000 | 15.00 |
| Ismaila et al (2016) | 35.00 | 4.50 | year | Infants | CHL | B | S | 1000 | 3.90 |
| Ismaila et al (2016) | 35.00 | 4.50 | year | Infants | CHL | B | M | 2000 | 14.00 |
| Ismaila et al (2016) | 35.00 | 4.50 | year | Infants | CHL | B | S | 2000 | 4.60 |
| Ismaila et al (2016) | 35.00 | 4.50 | year | Infants | CHL | B | M | 4000 | 13.50 |
| Ismaila et al (2016) | 35.00 | 4.50 | year | Infants | CHL | B | S | 4000 | 4.00 |
| Ismaila et al (2016) | 35.00 | 4.50 | year | Infants | CHL | A | M | 500 | 24.00 |
| Ismaila et al (2016) | 35.00 | 4.50 | year | Infants | CHL | A | S | 500 | 6.40 |
| Ismaila et al (2016) | 35.00 | 4.50 | year | Infants | CHL | A | M | 1000 | 23.50 |
| Ismaila et al (2016) | 35.00 | 4.50 | year | Infants | CHL | A | S | 1000 | 6.90 |
| Ismaila et al (2016) | 35.00 | 4.50 | year | Infants | CHL | A | M | 2000 | 23.50 |
| Ismaila et al (2016) | 35.00 | 4.50 | year | Infants | CHL | A | S | 2000 | 8.20 |
| Ismaila et al (2016) | 35.00 | 4.50 | year | Infants | CHL | A | M | 4000 | 22.50 |
| Ismaila et al (2016) | 35.00 | 4.50 | year | Infants | CHL | A | S | 4000 | 6.60 |
| Valeriote & Small (2024) | 23.00 | 7.36 | week | Infants | NH | A | M | 500 | 17.39 |
| Valeriote & Small (2024) | 23.00 | 7.36 | week | Infants | NH | A | S | 500 | 9.63 |
| Valeriote & Small (2024) | 21.00 | 7.36 | week | Infants | NH | A | M | 2000 | 21.00 |
| Valeriote & Small (2024) | 21.00 | 7.36 | week | Infants | NH | A | S | 2000 | 13.96 |
| Valeriote & Small (2024) | 15.00 | 6.71 | week | Infants | CHL | A | M | 500 | 15.33 |
| Valeriote & Small (2024) | 15.00 | 6.71 | week | Infants | CHL | A | S | 500 | 12.63 |

**Table S7:** Data. B = Behavioural BC Thresholds; A = ASSR BC Thresholds; M = Mean; S = SD; NH = Normal-hearing; SNHL = Sensorineural hearing loss; CHL = Conductive hearing loss; wk pca = post-conceptional age in weeks.
